# A systematic review of Flotation-Restricted Environmental Stimulation Therapy (REST)

**DOI:** 10.1101/2023.11.29.23299203

**Authors:** Elnaz Lashgari, Emma Chen, Jackson Gregory, Uri Maoz

**Affiliations:** Abbott Laboratories, Sylmar, CA, USA; Brain Institute, Chapman University, Irvine, CA, USA; University of California, Los Angeles, Los Angeles, CA, USA; California Institute of Technology, Pasadena, CA, USA

**Keywords:** Restricted Environmental Stimulation Therapy, float pod, flotation, PRISMA

## Abstract

**Background:** Restricted Environmental Stimulation Therapy (REST) is a therapeutic technique that involves immersing an individual in an environment with minimal sensory input or stimulation. The goal of REST is to induce a state of relaxation that is deeper than what can be achieved through other forms of relaxation techniques. Research suggests that REST can help reduce anxiety, alleviate chronic pain, improve sleep, and enhance creativity and cognitive function. Flotation-REST is a popular type of REST that utilizes an enclosed tank filled with buoyant saltwater to facilitate relaxation. This study aimed to synthesize the evidence on studies that investigate the effects of flotation-REST.

**Methods:** We used PRISMA to survey the flotation-REST literature from 1960 to 2023. This search was conducted on 29 January 2023 within the Google Scholar and PubMed databases. Journal and conference papers, as well as electronic preprints, that used flotation-REST in their methods, were written in English, and were published in or after 1960 were included; non-original research papers (e.g., review papers, book chapters, and papers solely on types of REST other than flotation-REST (e.g., chamber-REST) were excluded. From each eligible paper, we extracted information regarding the participant sample, application of flotation-REST, experimental design, treatment delivery method, questionnaires and tools, and study results.

**Results:** In total, we found 60 studies that included 1,838 participants. We propose that the application of flotation-REST can be divided into nine main categories: pain, athletic performance, physiology, stress, consciousness, psychology, creativity, clinical anxiety, sleep, smoking cessation, and other miscellaneous applications. In general, flotation-REST was found to bring about positive effects on pain, athletic performance, stress, mental well-being, and clinical anxiety, while having limited to no effect on sleep-related disorders and smoking cessation.

**Conclusion:** This paper provides a comprehensive overview of the current research on flotation-REST, highlights ongoing limitations in the literature, and outlines potential areas for future research. While flotation-REST appears to induce various benefits for physical and mental well-being, particularly when it comes to managing states like pain and stress, more research is needed to better understand the mechanisms underlying these effects and to identify optimal treatment protocols for different populations. A limitation of this paper is the relatively small number of studies available for review, which limits the generalizability of certain findings and highlights the need for additional research in this area.

## 1. Introduction

Restricted Environmental Stimulation Therapy (REST) is a therapeutic technique that involves immersing an individual in an environment with minimal sensory input or stimulation. Flotation-REST refers to a type of REST that involves floating in a tank filled with saltwater, also known as a float pod, float tank, or sensory deprivation tank. During flotation-REST, an individual is enclosed in a soundproof and lightproof chamber, floating in a shallow pool of water that is saturated with Epsom salt, which makes the water dense and enables the individual to float effortlessly. This environment is designed to minimize any sensory input from the outside world and thus to lead to a state of deep relaxation and reduced brain stimulation. The goal of all forms of REST is to induce a deep state of relaxation. As detailed below, previous research suggested that REST can help reduce anxiety, alleviate chronic pain, improve sleep, and enhance creativity and cognitive function. In clinical settings, REST has been proposed as a treatment for various conditions, including anxiety disorders, chronic pain, and post-traumatic stress disorder (PTSD). REST has also been used in sports therapy to help athletes recover from injuries and improve performance.

An early pioneer of flotation-REST research was John C. Lilly at The National Institute of Mental Health in the 1950s. It appears that Lilly created the first sensory deprivation tank in 1954 [1]. While Lilly played a significant role in popularizing the use of flotation tanks and exploring the concept of sensory deprivation in the 1950s and 1960s, it seems that he did not publish any specific scientific papers on the topic during that time. This early research on REST focused on elucidating the effects of reduced environmental stimulation on a range of psychophysiological, motoric, perceptual, cognitive, and emotional outcomes by placing people in sensory deprivation chambers for extended periods of time. The research eventually transitioned into an attempt to gain in-depth understanding of consciousness in the absence of external stimuli [2].

While Lilly was researching the effects of sensory deprivation though deep-water immersion, Donald Hebb investigated the effects of dry-REST, more commonly known as chamber-REST [3]. Chamber-REST occurs in a soundproof and lightproof room. The participant lies on a bed and is encouraged to limit movement. The experimental sessions tend to be long, lasting up to 24 (consecutive) hours. Researchers hypothesized that people were more prone to external influence and suggestion after periods of sensory deprivation; thus, panic buttons, medical trays, and other anxiety-provoking situational cues were sometimes introduced [4]. The view that sensory deprivation caused negative cognitive and consciousness effects persisted into the 1970s [5].

In 1968, Peter Suedfeld started conducting research on flotation-REST and eventually coined the term “Restricted Environmental Stimulation Therapy” (or REST). More extensive academic research then began globally, the majority taking place in the 2000s by Annette Kjellgren from Karlstads Universitet in Sweden [6–13]. In more recent years, Justin Feinstein and his team, then at the Laureate Institute of Brain Research in the United States, conducted the first functional magnetic resonance imaging (fMRI) study on the effects of flotation-REST [14].

Several review papers have been published on the effects of flotation-REST, focusing on either stress [15, 16] or sleep [17]. These papers and others provide insight into the potential benefits that have been claimed for flotation-REST, including reducing stress and anxiety, improving sleep and relaxation, and promoting general well-being. However, these previous reviews are relatively narrow in scope, discussing specific applications of flotation-REST What is more, to our knowledge, there has been no recent review of flotation-REST literature, despite progress in recent years. Hence, a more wholistic, comprehensive, and up-to-date review of flotation-REST comparing and contrasting its effects across different domains and applications is needed. This is the goal of this systematic review.

## 2. Methods

### 2.1. Search method for identification of relevant studies

This systematic literature review follows the preferred reporting items for systematic review and meta-analysis (PRISMA) guidelines [18]. PRISMA provides a transparent and structured framework for researchers to follow when conducting a systematic review, enhancing the transparency, reproducibility, and quality of the review. For the selection of potential articles to include in the review, the PRISMA guidelines recommend three phases: identification, screening, and inclusion.

During identification (Phase 1), we used keywords to search for potentially relevant papers. This search was conducted on 29 January 2023 within the Google Scholar and PubMed databases using the following group of keywords: “Restricted Environmental Stimulation Therapy” OR “Float pod” OR “Flotation REST” OR “Flotation pod” OR “Flotation Tank” OR “Sensory isolation”. The search yielded 2,439 papers (PubMed: 179 and Google Scholar: 2,260).

During screening (Phase 2), we screened the papers identified in Phase 1 for relevance. After reviewing the title and abstract of each paper, 2,146 papers were removed because they were not relevant, resulting in 293 remaining papers. We then further reviewed those 293 papers for duplicates; of those, 101 were duplicates. Thus, Phase 2 yielded 192 articles. The remaining 192 papers were examined in detail to meet the criteria set by the researchers.

During inclusion (Phase 3), authors EL and JG independently and manually reviewed the remaining papers to determine if they met the eligibility criteria for inclusion in this review (see Table 1). Screening the full texts of the remaining 192 papers suggested that 132 of them did not meet the eligibility criteria for this review (e.g., the relevant keywords were included in the references rather than in the paper itself, the paper waaas not published in English, the paper used dry chamber-REST only). Thus, we ended up with 60 papers that were included in this analysis (Figure 1).

**Figure 1.**
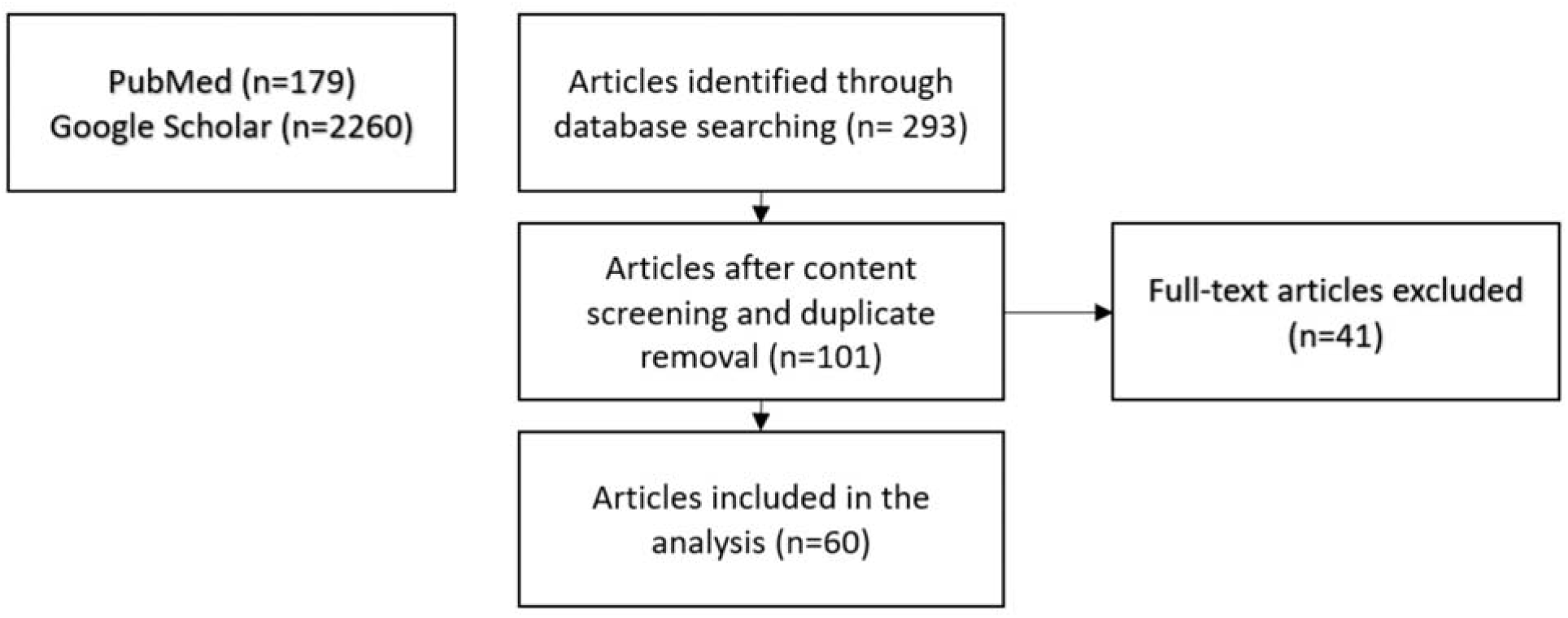
Selection process for the papers to include in the review process. See section 2.1 for details.

**Table 1.**
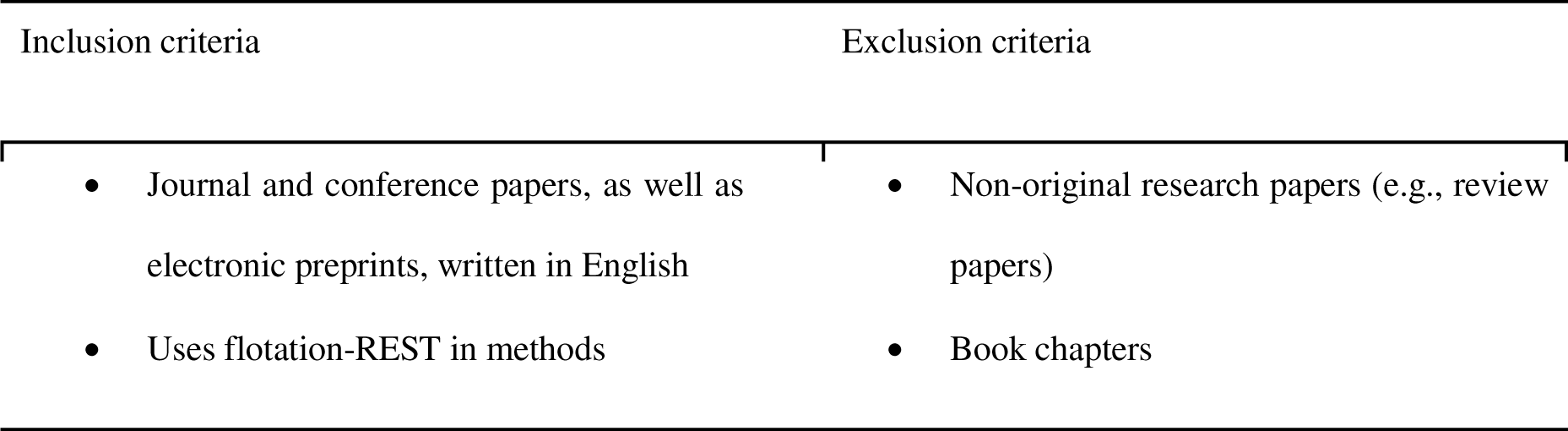
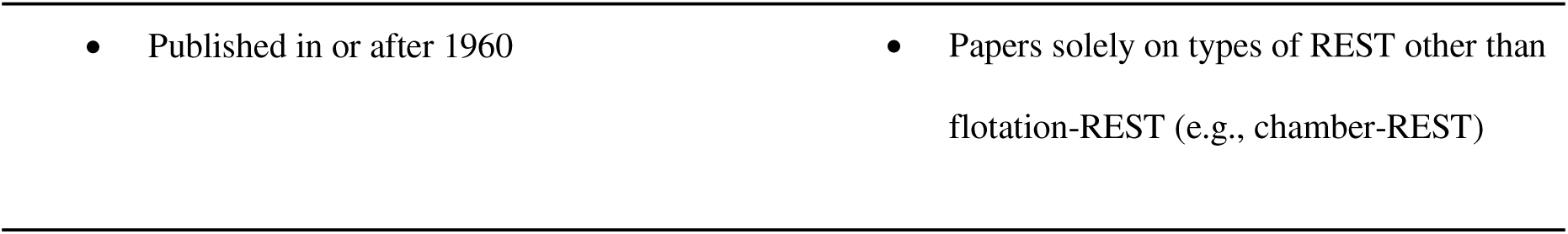
Predetermined inclusion and exclusion criteria for reviewed papers.

### 2.2. Data extraction and presentation

For each selected article, authors EL, JG, and EC independently extracted 30 features, covering categories: article origin; data sample; application of flotation-REST; experimental design; treatment delivery; questionnaires and tasks; and results (see Table 2 for details).

**Table 2.** Data items extracted for each article selected.

| Category | Data item |
| --- | --- |
| **Article origin** | Type of publication (journal article, conference article, or manuscript in an electronic preprint repository), location where the study took place |
| **Data sample** | Quantity of data, number of participants, age, selection method, conditions |
| **Application of flotation-REST** | Type of treatment carried out with flotation-REST |
| **Experimental design** | Description of randomization or pre/post design |
| **Treatment delivery** | Number of sessions, duration of sessions |
| **Questionnaires and tasks** | All the measurements taken during the study |
| **Results** | The effect of flotation-REST on the participants |

## 3. Results

### 3.1. Origin of the selected studies and different applications of float pod

The research methodology returned a total of 60 journal papers on flotation-REST. The majority of studies (44 out of 60, 73.3%) were conducted either in the United States (23 studies, 38.3%) or in Sweden (21 studies, 35.0%), with smaller numbers of studies conducted in Canada (8 studies, 13.3%), Japan (3 studies, 5.0%), Australia, Germany, Switzerland, the United Kingdom, and New Zealand (1 study each, 1.7%). These findings suggest that research on flotation-REST spans many countries, though it consists only of developed countries or members of the “global north”. The distribution of studies may also reflect differences in the availability and popularity of float pods in different regions, as well as varying levels of research interest and funding.

The reviewed papers were categorized into groups based on their application of flotation-REST (Figure 2). The following groups were identified: pain (10 studies), athletic performance (8 studies), physiological effects (8 studies), stress (6 studies), consciousness (5 studies), psychology (5 studies), creativity (5 studies), clinical anxiety (4 studies), sleep (3 studies), smoking cessation (2 studies), and other (4 studies). These categories reflect the diverse range of flotation-REST applications explored in the literature, highlighting the float pod’s utility across different fields of research.

**Figure 2.**
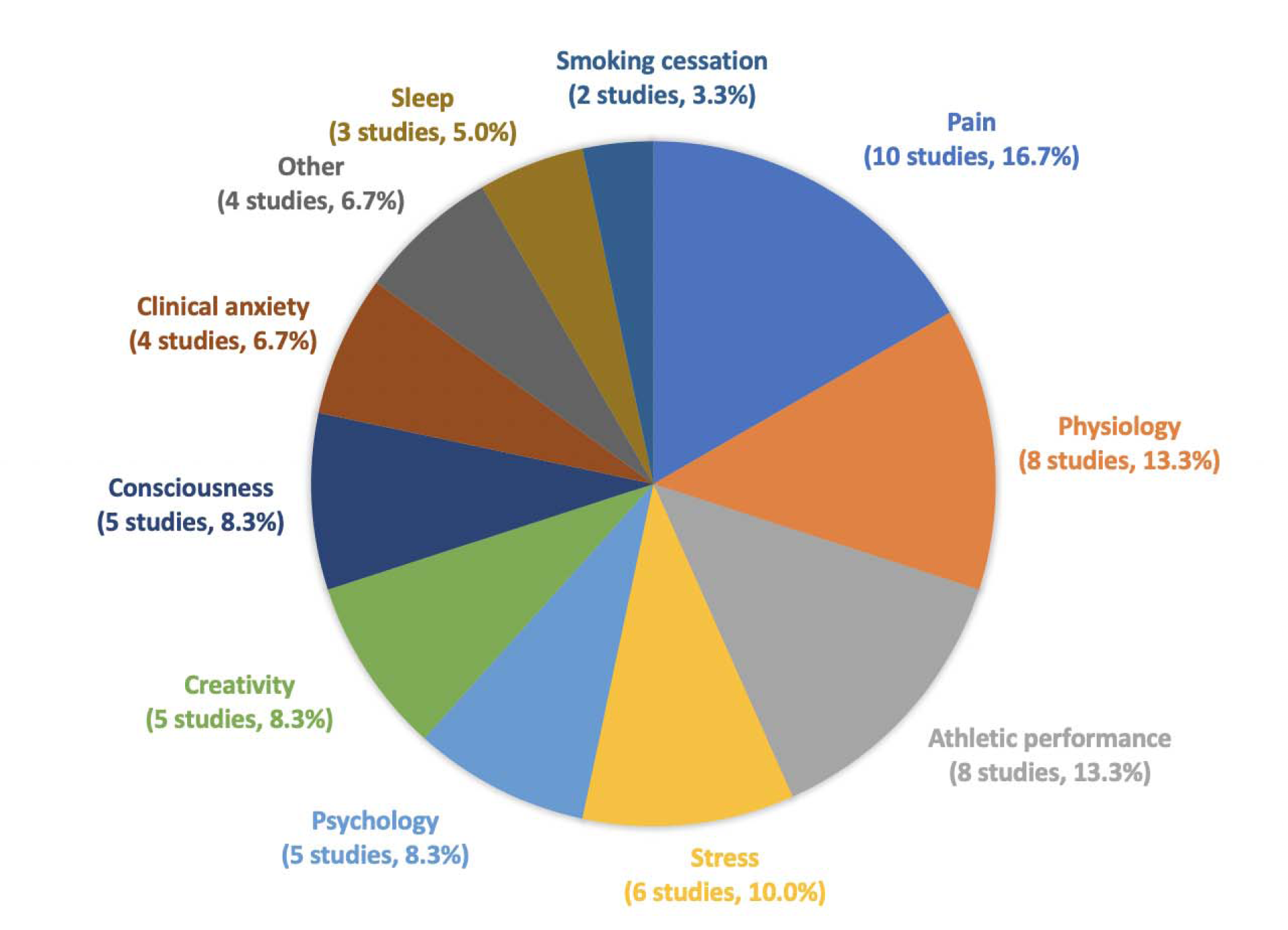
The percentage of flotation-REST studies in different flotation-REST applications across 60 reviewed studies. The following groups were identified: pain (16.7%, 10 studies), athletic performance (13.3%, 8 studies), physiology (13.3%, 8 studies), stress (10.0%, 6 studies), consciousness (8.3%, 5 studies), psychology (8.3%, 5 studies), creativity (8.3%, 5 studies), clinical anxiety (6.7%, 4 studies), sleep (5.0%, 3 studies), smoking cessation (3.3%, 2 studies), and other (6.7%, 4 studies).

### 3.2. Sample characteristics

The number of recruited participants in the 60 reviewed studies ranged from 1 to 99, with an average of 30.6±25.4 (mean ± standard deviation) participants per study. These findings suggest that research o flotation-REST is characterized by moderate sample sizes, similar to those common in neuroimaging studies. In total, the studies included 1,838 participants. Figure displays the overall number of participants in eac category. The pain category has the largest number of participants. The preponderance of participants in the pain category as well as that same pain category serving as the plurality among the percentages of studies among the different categories (Figure 2) suggests a special interest in float pods as tools for pain management. Another category receiving a lot of attention, relatively, is athletic performance (3^rd^ largest number of studies, Figure 2, and 2^nd^ largest number of participants, Figure 3).

**Figure 3.**
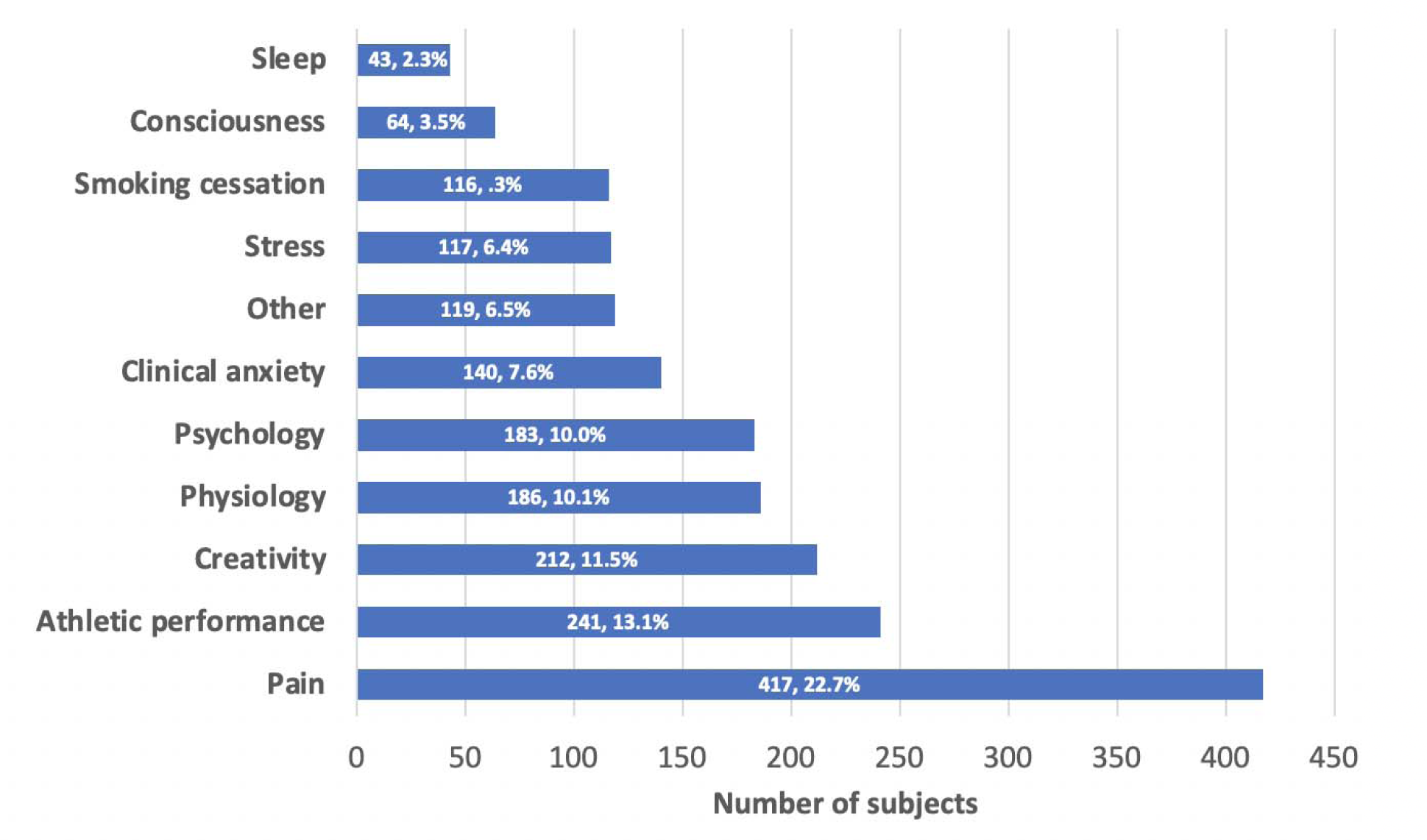
Total number of participants for different groups of flotation-REST and percentage across 60 reviewed studies

### 3.3. Experimental design

The reviewed studies used a variety of experimental designs (Figure 4). Three categories of experimental design were identified: (1) randomized designs, which included both within-group designs and randomize controlled trials (39 studies); (2) pre-test/post-test measurement design (11 studies); and (3) semi-structure interviews (7 studies). The remaining 3 studies did not fit into these categories. A range of experimental designs have therefore been used to investigate the effects of flotation-REST. The frequent use of randomized designs, i particular, reflects a rigorous research standard among flotation-REST studies. In contrast, pre/post measurement and semi-structured interview designs provide insight into the effects of flotation-REST in a more natural setting.

**Figure 4.**
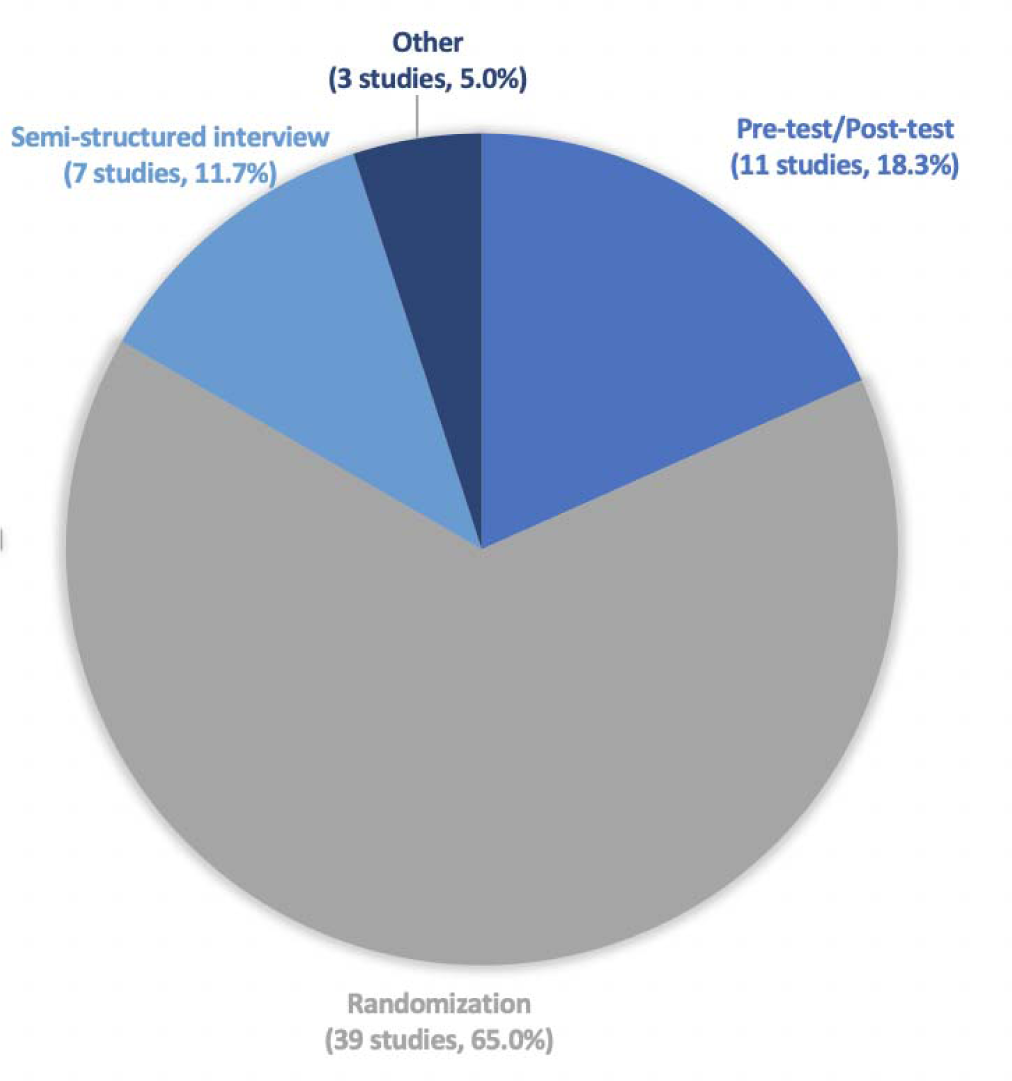
The percentage of experimental designs across 60 reviewed studies. Four categories of experimental design were identified: (1) randomization (65.0%, 39 studies); (2) pre-test/post-test measurement design (18.3%, 11 studies); (3) semi-structured interviews (11.7%, 7 studies), and (4) other (5.0%, 3 studies). In a randomization design, participants are randomly assigned to an experimental condition. In a pre-test/post-test design, participants complete assessments both before and after the experimental condition. Finally, in a semi-structured interview, participants respond to questions about their experience with the experimental condition.

### 3.4. Treatment delivery

The delivery of flotation-REST as treatment can vary depending on its intended therapeutic application. Here are some examples of how float pods have been used historically (see section 3.6 for more details):

- Pain management: Flotation-REST may help manage chronic pain by providing a sensory-deprived environment that reduces stimulation and promotes muscle relaxation. Treatment may involve a series of flotation sessions over a period of weeks or months.
- Athletic performance: Athletes recovering from injuries or seeking to enhance performance may benefit from flotation-REST, which may reduce muscle tension and promote relaxation. Treatment may involve a series of flotation sessions before or after a competition or be part of an athlete’s regular training program.
- Psychology and mental health: Float pod sessions may be used as a complementary therapy for a range of psychological and mental health conditions, such as anxiety, depression, and PTSD. Treatment may involve a series of flotation sessions over a period of weeks or months, combined with other therapies such as counseling and medication.
- Relaxation and stress reduction: Float pods may also be used in a nonclinical setting, to induce relaxation or to reduce stress in the general population, either through a single flotation session or a series of sessions over a period of time.

In general, flotation-REST typically involves a series of sessions over a period of time, with the number of sessions depending on the targeted therapy (Figure 5). The duration of each session can also vary and typically ranges from 35 to 90 minutes (Figure 6). Other therapies or interventions may be combined with flotation-REST to enhance its effects.

**Figure 5.**
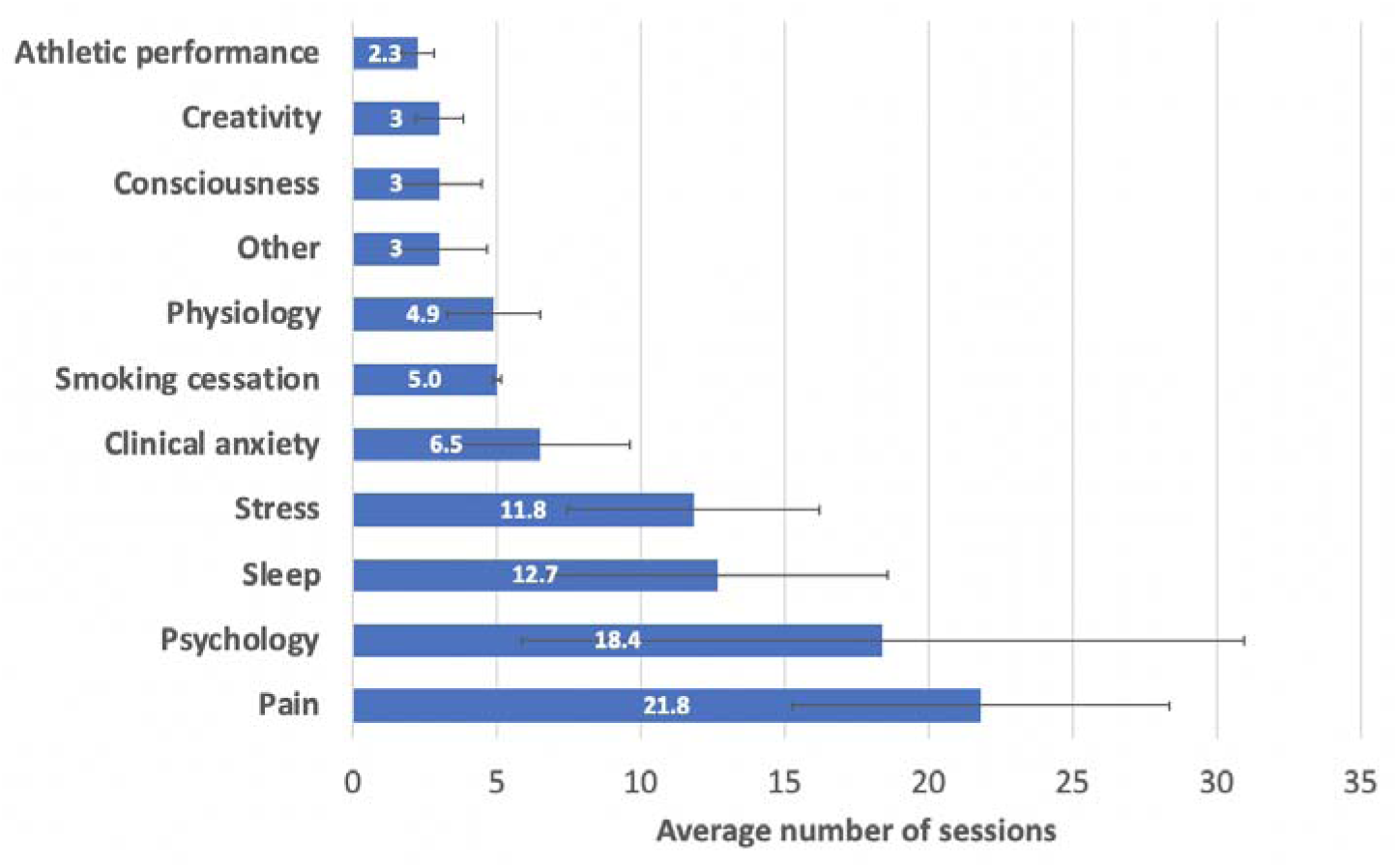
Average number of sessions for different groups of flotation-REST across 60 reviewed studies. Error bars represent 95% confidence intervals.

**Figure 6.**
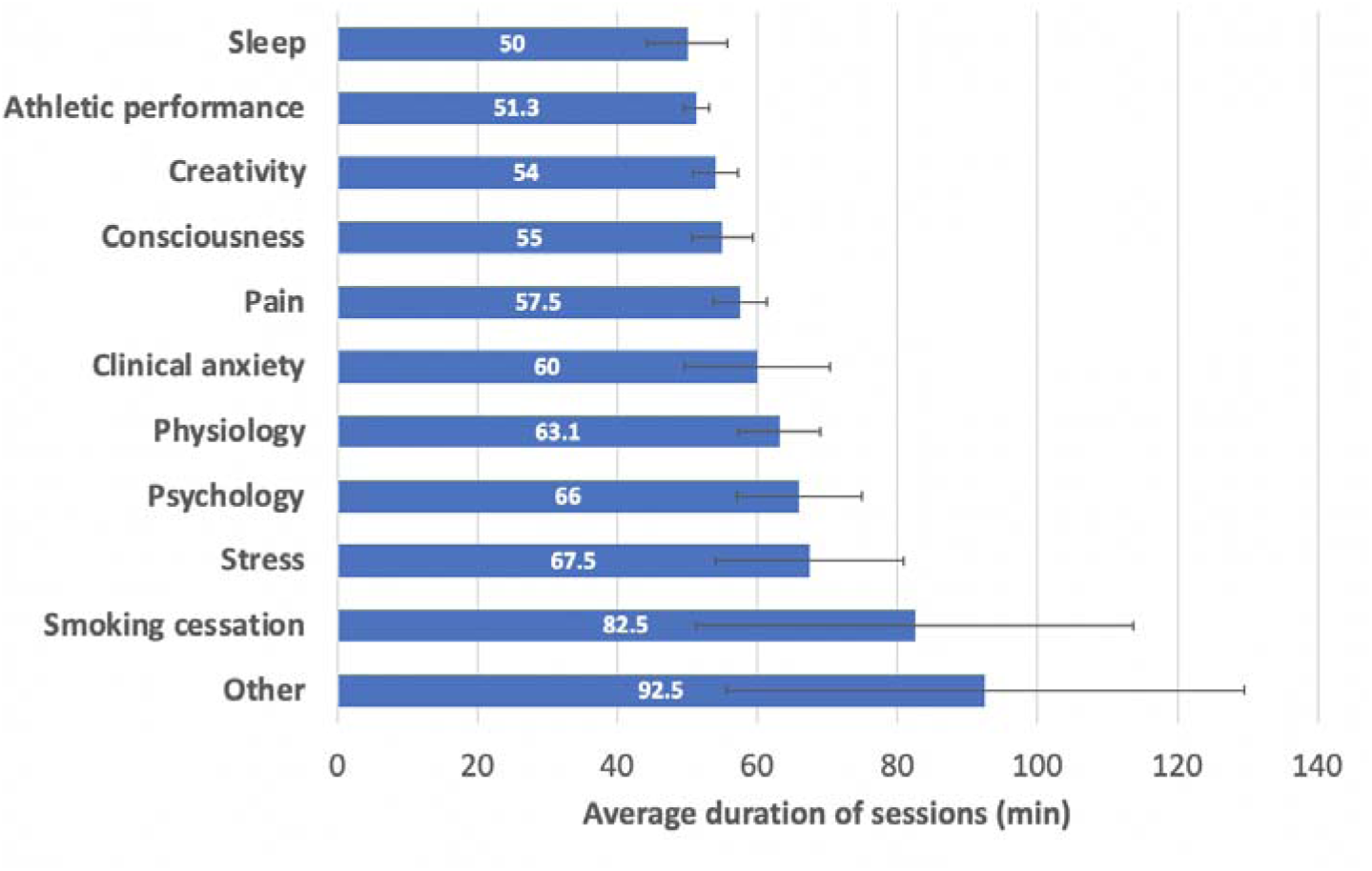
Average duration of sessions for different groups of flotation-REST across 60 reviewed studies. Error bars represent 95% confidence intervals.

**Figure 7.**
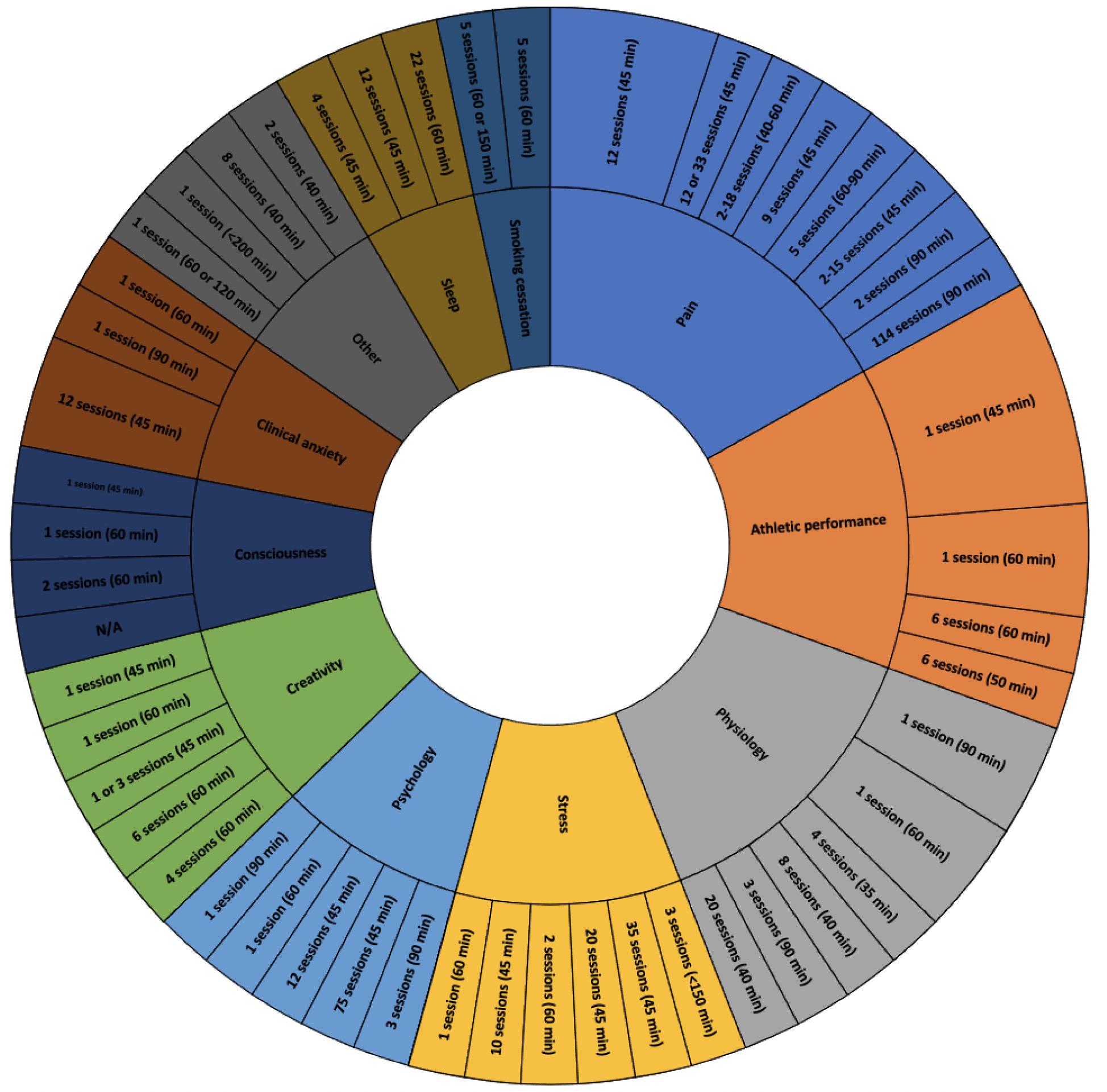
Treatment delivery (number of sessions and duration per session) for the different applications of flotation-REST (as in Figure 2) across the reviewed studies. Larger slices on the outer ring represent multiple studies that shared the same treatment delivery parameters.

Bayesian ANOVAs were performed to test for differences in the number and duration of sessions between different flotation-REST applications. These ANOVAs revealed that there is anecdotal evidence (BF_10_ = 0.353) in favor of there being no differences in the number of sessions between different flotation-REST applications and substantial evidence (BF_10_ = 0.159) in favor of there being no differences in the duration of sessions between different flotation-REST applications.

### 3.5. Questionnaires

Questionnaires are commonly used in flotation-REST research to assess various outcomes, such as pain, stress, anxiety, mood, and cognitive performance. Questionnaires are a useful tool for assessing subjective experiences and outcomes in flotation-REST, and a range of validated questionnaires have been used to assess different outcomes. In addition to questionnaires, other measures such as heart rate, blood pressure, cortisol levels, and electroencephalography (EEG) have been used in flotation-REST research to assess various physiological and neurobiological outcomes. Table 3 shows the list of all tasks and questionnaires used as outcome measures (i.e., not for mere screening purposes) in the 60 studies. As is apparent from the table, the most common questionnaires were the HADS, PANAS/PANAS-X, and POMS, which were used as outcome measures across a total of 7 studies each. Other popular questionnaires included the STAI (used in 6 studies); EDN, LOT, SE, and VAS (5 studies); and PAI (4 studies). Additionally, 13 studies used unique questionnaires (i.e., original questionnaires that were made for the purpose of the study) while 21 studies did not use any questionnaire at all as an outcome measure.

**Table 3.** Questionnaires and tasks used in the studies included in this review paper. References to the studies in which the test was used are provided in the last column of the table. In the second column, we denote any tests that are a task rather than a questionnaire with “(task)”.

| Test name | Description | Number of times used |
| --- | --- | --- |
| **AST** | Arousal Seeking Tendency | 1 [15] |
| **BAS-2** | Body Appreciation Scale 2 | 1 [66] |
| **BCS** | Body Consciousness Scale | 1 [15] |
| **BDI** | Beck Depression Inventory | 2 [33, 74] |
| **Beer Can/Brick** | A test of divergent thinking that measures the number of relevant responses for how many different ways one may use a beer can or a brick, respectively. (task) | 1 [70] |
| **BISS** | Body Image States Scale | 1 [66] |
| **CSAI-2** | Competitive State Anxiety Inventory-2 | 1 [41] |
| **DERS** | Dysfunctional Emotional Regulation Scale | 1 [6] |
| **EDEQ-6** | Eating Disorder Examination Questionnaire 6 | 1 [66] |
| **EDN** | Experienced Deviation from Normal state scale | 5 [6, 12, 13, 27, 70] |
| **Frego Test** | A test that measure “preconscious activity”. (task) | 1 [68] |
| **FS – Change and Stability Test** | A test that measures attitude to change and stability. (task) | 2 [68, 70] |
| **GAD-Q-IV** | Generalized Anxiety Disorder Questionnaire (4th Edition) | 1 [6] |
| **GCS** | Guilford Creativity Scale | 1 [5] |
| **HADS** | Hospital Anxiety and Depression Scales | 7 [13, 25, 26, 27, 28, 31, 70] |
| **HPT** | Haptic Processing Test (task) | 1 [46] |
| **IAM** | Interoceptive Awareness Measure | 1 [66] |
| **ISI** | Insomnia Severity Index | 1 [76] |
| **ISQ** | Isolation Symptom Questionnaire | 1 [57] |
| **KSS** | Karolinska Sleep Scale | 1 [72] |
| **LOT** | Life Orientation Test | 5 [13, 25, 26, 28, 70] |
| **MAACL** | Multiple Affect Adjective Check List | 1 [57] |
| **MAAS** | Mindful Attention Awareness Scale | 2 [6, 13] |
| **MADRS-S** | Montgomery-Asberg Depression Rating Scale | 2 [6, 76] |
| **MDMQ** | Multidimensional Mood Questionnaire | 1 [40] |
| **MMPI** | Minnesota Multiphasic Personality Inventory | 1 [47] |
| **Muscle Soreness** | A scale ranging from 0 (not sore) to 10 (maximum soreness). | 1 [40] |
| **PAI** | Pain Area Inventory | 4 [26, 27, 28, 31] |
| **PANAS/PANAS-X** | Positive and Negative Effect Schedule (Expanded Form) | 7 [26, 27, 28, 54, 66, 72, 73] |
| **PDI** | Pain Disability Index | 1 [33] |
| **PE** | Pain Endurance (task) | 1 [31] |
| **PEQ** | Performance Evaluation Questionnaire | 1 [36] |
| **PFRS** | Photographic Figure Rating Scale | 1 [66] |
| **POMS** | Profile of Mood States | 7 [5, 58, 63, 64, 67, 74, 79] |
| **PSQI** | Pittsburgh Sleep Quality Index | 1 [6] |
| **RMS-Person/RMS-Place** | Russell Mood Scale-Person/Russell Mood Scale-Place [19]<br>[20] | 1 [15] |
| **RPE** | Ratings of Perceived Exertion | 1 [39] |
| **SAS** | State Anxiety Scale | 1 [57] |
| **SE** | Stress and Energy | 5 [13, 26, 27, 28, 31] |
| **SFHS** | Short Form Health Survey | 1 [33] |
| **SPI** | State Personality Inventory | 3 [5, 58, 63] |
| **SQ** | Sleep Quality | 2 [13, 33] |
| **SRAS** | Self-Report Arousal Scale | 1 [74] |
| **SRQ** | Subjective Relaxation Questionnaire | 1 [56] |
| **SSS** | Subjective Stress Scale | 1 [47] |
| **SSS-IV** | Sensation Seeking Scale – Form IV [21] | 1 [47] |
| **STAI** | State-Trait Anxiety Inventory | 6 [33, 43, 54, 66, 72, 73] |
| **Syllogisms I-II** | A test presented in two versions [36] that measures the ability of logical and deductive thinking. (task) | 2 [68, 70] |
| **VAS** | Visual Analogue Scales | 5 [13, 26, 47, 72, 73] |
| **WBPS** | Wong-Baker Pain Scale | 1 [72] |
| **WPI** | Widespread Pain Index | 1 [33] |
| **WRMT** | Warrington Recognition Memory Test (task) | 1 [46] |
| **ZMQ** | Von Zerssen's Mood Questionnaire | 1 [47] |

On average, 1.9±1.3 questionnaires were used per study. Studies in the psychology category had the highest average of 3.6±3.6 questionnaires per study, followed by clinical anxiety (3.5±2.6), pain (3.2±2.6), creativity (2.8±2.7), sleep (2.3±2.1), stress (1.7±1.6), physiology (1.5±2.0), athletic performance (0.8±0.7), consciousness (0.6±0.9), and other (0.5±0.6). No study in the smoking category used any questionnaire.

### 3.6. Float pod applications

We categorized the reviewed papers into various groups based on their application of flotation-REST (Figures 2 and 5). Here is a brief summary of the main findings from each study category:

- Pain (10 studies): Flotation-REST can effectively reduce pain stemming from a variety of conditions, including chronic tension headaches, stress-induced muscle tension, and whiplash associated disorder.
- Athletic performance (8 studies): Flotation-REST may improve certain components (e.g., accuracy, precision) of athletic performance, especially when coupled with guided imagery tasks, as well as enhance performance recovery.
- Physiology (8 studies): Flotation-REST was generally found to have various physiological effects indicative of decreased sympathetic arousal, such as reduced blood pressure, slowed breathing rate, and decreased cortisol levels.
- Stress (6 studies): Flotation-REST was found to reduce levels of stress in both clinical and non-clinical participant samples.
- Consciousness (5 studies): Altered states of consciousness are frequently induced by flotation-REST, with participants reporting experiencing visual and auditory hallucinations, distorted time perception, out-of-body experiences, and personally profound and transformative visions.
- Psychology (5 studies): Across both healthy and non-healthy participants, flotation-REST was found to induce positive effects on various psychological outcomes, including improved mood and reduced anxiety and depression.
- Creativity (5 studies): In general, flotation-REST was found to enhance certain facets of creativity, such as originality, divergent thinking, and technical musical ability.
- Clinical anxiety (4 studies): Among participants with a wide range of anxiety-related disorders, flotation-REST was found to produce anxiolytic effects.
- Sleep (3 studies): Flotation-REST was found to have limited potential as a treatment for sleep-related disorders (e.g., insomnia). Two studies found evidence of flotation-REST improving sleep latency, but one of these studies found that this improvement only reached significant levels three months after treatment completion, while the other study found that only half of the participants experienced improvement.
- Smoking cessation (2 studies): Compared to other, more established therapeutic interventions for smoking cessation, flotation-REST is not particularly effective in helping individuals reduce their smoking habits.
- Others (4 studies): The remaining studies explored components of flotation-REST protocol (2 studies), sought to investigate hypotheses about flotation-REST (1 study), or examined miscellaneous effects of flotation-REST (1 study).

Overall, the reviewed studies suggest that flotation-REST can have various benefits for physical and mental health, but more research is needed to better understand the mechanisms underlying these effects and to identify optimal treatment protocols for different populations.

Below we review the flotation-REST protocol, methods, and findings for each study, categorized by application. We hope that this review will be valuable for future flotation-REST research by serving as a reference for experimental design, helping elucidate existing gaps in knowledge, and inspiring new research directions.

#### 3.6.1. Pain

We found ten studies that investigated the effects of flotation-REST on pain. Based on these studies, flotation-REST appears to be an effective method for pain relief. All ten studies found evidence in support of the analgesic effects of flotation-REST; notably, flotation-REST was found to alleviate pain stemming from a variety of disorders, including stress-related muscle tension (5 studies), WAD (2 studies), chronic pain disorder (2 studies), and tension headaches (1 study). Importantly, however, the only study in this category to compare flotation-REST to placebo found comparable results between the two treatments [33]. Out of the eight quantitative studies in this category, all studies (8 out of 8; 100.0%) found significant results in favor of flotation-REST reducing pain. If we assume that the probability of a study finding a significant result is 50% (i.e., a coin toss), a Bayesian binomial test shows that there is very strong evidence (BF_+0_ = 56.778) in support of the hypothesis that the likelihood of a study finding that flotation-REST can significantly improve pain is greater than chance. Given this and the high number of studies in this category relative to the other categories, we can reasonably assume that flotation-REST is capable of relieving pain across multiple disorders, although whether this benefit is partially due to the placebo effect remains unclear. Below, we provide a summary for each of the ten studies.

In 1985, Fine and Turner sought to assess the effects of flotation-REST on chronic pain [22]. 15 participants diagnosed with chronic pain disorder completed a multimodal treatment program that included psychotherapy, EMG-based biofeedback training, and flotation-REST. Participants completed an average of 13 25-minute biofeedback training sessions and an average of 7 flotation-REST sessions ranging from 40 to 60 minutes in length. Each participant reported their pain intensity, frequency, and duration before and after completion of the treatment program. In a follow-up interview at least three months post-treatment, participants were also asked to compare the depth of relaxation achieved by each program mode. The results showed that participants reported a small but significant decrease in pain intensity (but not frequency or duration) following the completion of the treatment program. Additionally, flotation-REST was found to be significantly more relaxing than the EMG-based biofeedback training. The authors suggest that flotation-REST may be a promising therapeutic intervention for chronic pain because of its ability to induce deep relaxation.

Wallbaum et al. (1991) investigated the treatment potential of flotation-REST in individuals with chronic tension headaches [23]. 31 participants diagnosed with tension headache were randomly assigned to participate in one of four treatment conditions: 1) Chamber only (control condition), in which all study sessions were conducted in a small, quiet, dimly lit room where participants laid supine on a bed; 2) Chamber/tank, in which participants quietly rested in the control room for half their sessions and floated in a flotation tank for the other half; 3) Chamber/relaxation, in which participants quietly rested in the control room for half their sessions and completed progressive muscle relaxation exercises in the control room for the other half; and 4) Tank/relaxation, in which participants engaged in both flotation-REST and the muscle relaxation exercises. Each condition involved two 90-minute sessions per week for four weeks. A headache diary [24] was given to participants to track the frequency, duration, and intensity of their headaches, and these entries were then used to calculate a headache index for each participant. Participants completed entries in the diaries from at least two weeks before the first treatment until 6 months after completion of the study. Participants also reported any headache-related medication use before and during the study. The results indicated that all four conditions led to a significant decrease in the average mean headache index score over time. However, participants in the three active treatment (i.e., non-control) conditions demonstrated maintained improvement in their headache symptoms at the 6-month follow-up mark that was absent in the control group. Based on their findings, the authors concluded that flotation-REST may serve as an effective treatment for chronic tension headaches, and the duration of these therapeutic effects may be increased by combining flotation-REST with muscle relaxation exercises [23].

Kjellgren et al. (2001) investigated whether flotation-REST might serve as a therapeutic tool for alleviating chronic muscle pain [25]. 37 participants who reported experiencing muscle tension pain in the neck and back were recruited and were randomly assigned to a control group (no treatment) or an experimental group involving nine flotation-REST sessions carried out over three weeks. Each flotation session lasted 45 minutes, resulting in a total of 300 hours of treatment for each participant in the experimental group. At the start and end of the study, participants completed several measures, including ratings of their pain severity at its worst and their average pain severity, duration, and onset; subjective sleep quality; HADS; LOT; and blood samples. The results indicated that, on average, participants experienced a significant decrease in both average pain severity and pain severity at its worst. Furthermore, circulating levels of the noradrenaline metabolite 3-methoxy-4hydroxyphenylethyleneglycol were significantly reduced in the float-REST treatment group but not in the control group, whereas endorphin levels were not affected by flotation. Flotation-REST treatment also elevated the participants’ optimism while reducing anxiety or depression. Finally, participants in the flotation-REST treatment group reported being able to fall asleep more easily compared to those in the control group. The present findings tentatively suggest that, among other health benefits, flotation-REST may effectively alleviate low to moderately severe pain induced by muscle tension.

In 2005, Bood et al. examined the therapeutic effects of float-REST for stress-related muscle pain while also assessing if the degree of attention participants received during treatment might modulate the therapeutic effects by acting as a placebo [26]. 32 participants who had been diagnosed with stress-related muscle tension were recruited and randomly assigned to either receive special attention throughout the experiment for 12 weeks (high attention group) or receive normal attention for only six weeks (normal attention group). Those in the high attention group met and interacted with the laboratory staff on 24 occasions, double the number of that of the normal attention group. Both groups, however, completed 12 flotation sessions lasting 45 minutes each. The study used a pretest-posttest design; participants completed the PAI to assess features of their pain (intensity, areas and types, frequency, duration, onset and treatment efficacy) along with the PANAS, LOT, SE and HADS questionnaires. The results demonstrated that, across both groups, participants significantly exhibited lowered blood pressure; reduced pain, anxiety, depression, stress and negative affect; and increased optimism, energy and positive affect. Notably, the results were largely unaffected by the degree of attention received. The authors concluded flotation-REST may be an effective and non-invasive method for alleviating stress-related muscle pain, and that flotation-REST is not particularly sensitive to attention-placebo more so than other forms of pain therapy.

Bood et al. (2006) investigated the long-term effects of flotation-REST on stress-related muscle tension pain [27]. Seventy participants were recruited, with all participants having been diagnosed with muscle tension pain; 26 of these participants also were diagnosed with depression. Participants were randomly assigned to either a control group or a flotation–REST group, and each group completed a total of 12 45-minute sessions. These sessions occurred approximately twice a week over a 7-week period. Those in the control group rested in an armchair for their sessions. Before and after the treatment program, participants completed blood samples along with a questionnaire that included the PAI, SE, HADS, PANAS, EDN measures as well as a VAS for sleep quality. Results from before and immediately after the treatment program indicated that there were significant differences between the flotation-REST group and the control group, such that those in the flotation-REST group experienced lower levels of pain, stress, anxiety, and depression and greater levels of sleep quality, optimism, and prolactin. Additionally, analyses of the flotation-REST group 4 months after completion of the treatment program showed that these positive effects persisted with the exception of changes in prolactin levels. The authors conclude that flotation-REST may yield long-term physical and psychological health benefits for those with stress-related muscle tension pain.

To investigate whether longer treatment programs might increase the effectiveness of flotation-REST for stress-related muscle pain, Bood et al. (2007) compared the therapeutic effects of 33 flotation sessions to 12 sessions [28]. 37 participants who had been diagnosed with stress-related muscle tension were recruited and randomly assigned to the 12-session or 33-session treatment programs. Flotation-REST sessions across both groups were 45 minutes in duration. Before and after completion of the treatment program, participants completed a questionnaire comprised of the PAI, SE, HADS, LOT, and PANAS. Participants also had their blood pressure taken completed a test in which their pain levels were assessed by a pain matcher device. The results after 12 sessions were similar to those found in other float-REST studies [21]; surprisingly, there were little to no differences in treatment effects between the participants in the 33-session group and those in the 12-session group. Participants in both groups reported similar improvements in most severe pain intensity, normal pain intensity, or pain frequency, as well as similar levels of decreased stress, anxiety, negative affect, and depression and increased dispositional optimism and sleep quality. The only measures that significantly differed between the two groups were the number of comprehensive pain areas and diastolic blood pressure, which were both significantly lower after 33 flotation sessions but not 12 sessions. These findings suggest that flotation-REST on its own may not yield additional benefits for stress-related muscle tension beyond 12 treatment sessions.

In 2008, Edebol et al. investigated whether flotation-REST might improve pain-related symptoms stemming from chronic whiplash associated disorders (WAD) [29]. Six women and one man were recruited for the study. All participants had been diagnosed with chronic WAD and had experience with flotation-REST treatment for their condition, albeit to varying degrees (participants had completed a range of 2 to 15 flotation sessions). Participants completed a semi-structured qualitative interview during which they shared their experience with chronic WAD and flotation-REST treatment for their condition. The Empirical Phenomological Psychological (EPP) method based on [30] was then used to analyze the interviews. Based on the experiences of each participant, the authors identified five distinct stages that occur during flotation-REST treatment sessions for chronic WAD: (1) intensification, in which pain initially increases; (2) vitalization, in which pain and stress gradually decrease while relaxation increases; (3) transcendence, in which altered states of consciousness are typically experienced; (4) defocusation, in which participants arrives at new insights on how to manage their pain; and (5) reorientation, in which participants integrate these insights to develop a more optimistic perspective on their condition. The authors conclude that flotation-REST may improve both the physiological and cognitive symptoms associated with chronic WAD, although they note that qualitative data collected from more participants is necessary to validate this initial finding.

Building on their previous work investigating the effects of float-REST for stress-related muscle pain, Bood et al. (2009) sought to explore if sex differences might modulate these effects [31]. 88 participants were recruited and completed a similar protocol to [27] and [28], during which they completed 12 45-minute sessions over 7 weeks. The results indicated that there were few sex differences in the beneficial effects of flotation-REST treatment, which included decreased pain (which was significantly reduced), stress, anxiety, and depression and increased sleep quality. Notably, however, female participants reported higher depression scores than male participants before the flotation-REST treatment, whereas this difference was absent after treatment, suggesting that female participants experienced greater improvements in depression. Additionally, there was a sex difference in the ability to endure experimentally induced pain, such that men exhibited greater endurance both before and after the flotation-REST treatment. However, both sexes improved their ability to endure experimentally induced pain following the completion of their flotation-REST treatment. The authors conclude that flotation-REST can be recommended to both men and women as a treatment for stress-related muscle pain.

Similar to their 2008 study [23], Edebol et al. 2013 sought to evaluate the benefits of flotation-REST for chronic WAD. In this study, a single participant was interviewed. The participant had been diagnosed with chronic WAD - grade IV, indicating an especially severe case of WAD, and had been using flotation-REST to treat his condition for a year and a half, during which he recorded his experiences in a diary. A semi-structured interview was also conducted at the end of his flotation-REST treatment. for the first time, examined experience from long-term flotation REST made by a patient with chronic Whiplash Associated Disorder (WAD), grade IV [32]. The patient of the present study was a middle aged native-born Caucasian male from Sweden who had been diagnosed with chronic WAD IV by a licensed physician. The patient performed regular flotation for one and a half year and wrote about his experiences in a diary. A semi-structured interview was conducted at the end of therapy. Using the EPP method, the diary and interview were analyzed, and the findings indicated that participant experienced a host of positive effects from flotation-REST, including pain relief, increased relaxation, better mental coping, enhanced energy, muscular and motor improvements and increased subjective well-being. The authors conclude that these findings suggest that flotation-REST may holistically enhance the quality of life even for patients with severe chronic WAD.

In 2021, Loose et al. directly compared the effects of flotation-REST to an indistinguishable placebo for alleviating chronic pain [33]. 99 participants who had been diagnosed with chronic pain were recruited and randomly assigned to one of three groups: (1) a wait-list control group, (2) a placebo group, and (3) a flotation-REST group. Participants in the intervention and placebo groups underwent 5 treatment sessions lasting 60 to 90 minutes over the course of three weeks. Participants in the placebo group completed their sessions in a modified floating tank with reduced buoyancy and fewer environmental stimulus restrictions. All participants completed a series of measures at baseline and follow-ups assessments that evaluated pain intensity, pain-related disability (PDI), pain area, pain widespreadness (WPI), anxiety (STAI), depression (BDI-II), quality of life (SFHS), sleep quality, and use of pain medication. The results indicate that participants in the flotation-REST group experienced no long-term benefits (i.e., benefits lasting past the 24-week mark after the last treatment session) from the intervention. Additionally, short-term significant improvements in pain intensity, relaxation, anxiety, pain area, and pain widespreadness were found in similar degrees between the placebo and intervention groups, suggesting that previous findings indicating that flotation-REST may be an effective therapy for alleviating pain can be explained in part by placebo effect.

#### 3.6.2. Athletic performance

We found eight studies that investigated the effects of flotation-REST on athletic performance. Based on these studies, flotation-REST may be a useful therapeutic intervention for athletes. Seven out of eight studies found evidence in support of this conclusion; five of these positive studies found that flotation-REST, often coupled with an imagery task, can improve certain types of athletic performance, and the remaining two studies found that flotation-REST can enhance post-athletic performance recovery. Seven out of eight studies (87.5%) found significant results in favor of flotation-REST’s athletic-related benefits. If we assume that the probability of a study finding a significant result is 50% (i.e., a coin toss), a Bayesian binomial test shows that there is moderate evidence (BF_+0_ = 6.972) in support of the hypothesis that the likelihood of a study finding that flotation-REST can produce athletic-related benefits is greater than chance. Given this and the high number of studies in this category relative to the other categories, we can reasonably assume that flotation-REST may help athletes improve and recover from their athletic performance. We provide summaries for each of the eight studies below.

In 1990, Suedfeld and Bruno investigated the effect of flotation-REST, coupled with a guided mental imagery task, on athletic performance [34]. 30 participants were recruited, all of whom had little to no experience playing basketball and were divided into 3 treatment groups: flotation-REST, alpha chair, and control. Each participant completed a single 1-hour treatment session; before the session, all participants were asked to shoot 20 regular free throws. During their treatment sessions, those in the flotation-REST group floated in a dark, soundproofed tank; those in the alpha chair condition rested in a shell-like chair which enclosed the participant and was specifically “designed to induce relaxation and concentration”; and those in the control group sat in a regular comfortable armchair situated within an office. Then, while in their assigned environments, participants listened to an hour-long tape recording that directed them through a multisensory imagery task in which participants imagined themselves making basketball free throws. Finally, a day after the treatment session, participants completed an additional 20 free throws. Questionnaires were administered to assess previous athletic and guided imagery experiences; participants’ confidence in their athletic ability was also measured using self-reported predictions of their own free-throw success. The results showed that participants in the flotation-REST group made significantly more successful free throws than participants in either the alpha chair or control groups. Additionally, those in the flotation-REST group reported higher post-treatment levels of confidence in athletic performance compared to the alpha chair group (but not the control group). The authors conclude that their findings “provide strong empirical evidence of improvement in an athletic skill after flotation REST.”

McAleney et al. (1990) investigated the joint effects of flotation-REST and a visual imagery task on the competitive performance of intercollegiate tennis players [35]. The study sample consisted of 20 university varsity tennis players (10 men and 10 women), and participants were randomly assigned into one of two conditions: flotation-REST with the visual imagery task, or a visual imagery-only control condition in which participants completed the visual-imagery task in a REST chamber. The visual imagery task was eight minutes in length and included five or six images during which the participants visualized themselves making optimal tennis shots. All participants participated in six 50-minute treatment sessions across a 3-week period. To measure improvement in athletic performance, tennis matches from both before and after completion of the treatment sessions were videotaped for each participant. Performance was then assessed based on three key aspects: first service, key shot (i.e., the shot that resulted in winning or losing the game point), and points won/lost. The results indicated that participants in the flotation-REST group performed significantly better than participants in the control group only for the measure of first service accuracy. Based on these findings, the authors suggest that flotation-REST may be effective for enhancing highly controlled skills like the service motion but not for more variable movements like the key shot.

In 1991, Wagaman et al. investigated the joint effects of REST and an imagery task on subjective and objective measures of athletic performance in basketball [36]. 22 male basketball players were randomly assigned to participate in either REST-imagery or an imagery-only treatment. Participants completed six sessions of their assigned treatment over a five-week period. During each 60-minute session, those assigned to the REST-imagery treatment rested in a supine position within a float tank, while those assigned to the imagery-only treatment sat in a comfortable chair situated within an office where they were free to do as they pleased once a tape relaying the imagery training task had finished playing. The imagery task consisted of a 20-minute tape that guided the listener to relax and visualize the steps of an optimal basketball game performance. An objective measure of athletic performance was made using a point-based system assessing participants’ individual performances during real basketball games. Successful shots or passes resulted in participants gaining a point, whereas unsuccessful shots or passes resulted in participants losing a point. Subjective measures of athletic performance were also collected using the PEQ [37], a short survey filled out by the participants’ coaches who were blind to the participants’ treatment assignment. The results showed that the REST-imagery group’s athletic performance was significantly better than the imagery-only group on both objective point-based score and coaches’ subjective ratings of passing and shooting. The authors conclude that flotation-REST appears to increase the effectiveness of imagery training in boosting athletic performance.

In 1993, Suedfeld et al. examined the effects of the combination of flotation-REST with imagery training on perceptual-motor accuracy in comparison to flotation-REST and imagery training alone [38]. 40 participants who had played a game of darts at least twice during the previous year were recruited and randomly assigned to one of four groups: imagery only, flotation-REST only, flotation-REST plus imagery, and a control group. Prior to completing their assigned treatment, participants completed 20 recorded dart throws at a standard dart board. Then, those in the imagery-only group sat in a small room, reading or studying. After 40 minutes, participants were asked to listen to a 13-minute tape recording that started with a brief exercise during which participants were asked to describe the “feel” of throwing a perfect bull’s eye. Participants in the flotation-REST only group floated in a supine position within a flotation tank for 1 hour without any imagery training. Participants in the flotation-REST plus imagery group also rested in a supine position within a flotation tank but, after 40 minutes had elapsed, also completed the imagery training. Lastly, participants in the control group were placed within a small room and were told they could do as they pleased for the entire 1-hour session. Following their treatment session, participants completed another 20 recorded dart throws. Participants were scored based on the distance of their dart from the bullseye on the dart board. The results indicated that there was a significant main effect of flotation-REST on posttest scores; imagery training alone did not increase dart-throwing accuracy, and no interaction between flotation-REST and imagery training was found. From this, the authors concluded that flotation-REST may improve accuracy and precision in athletic performance for sports that do not require speed or strength.

Norlander et al. (1999) investigated the effect of flotation-REST on archery performance in archers of varying skill levels [39]. 20 participants were recruited and divided into three groups based on their adjudged skill (beginner, intermediate, or elite) by expert archery coaches. Participants were then randomly assigned to complete one 45-minute session of either flotation-REST or a control session that involved sitting in an armchair. Following the session, participants completed an archery performance task in which they were required to shoot a total of 12 arrows at fixed target, with the goal of hitting the target as close to its bullseye as possible. During this task, EMG recordings were obtained to determine muscle tension in the arms. After completing the task, participants then rated their estimated perceived exertion using the RPE scale. The results showed that those in the flotation-REST group experienced significantly less perceived exertion following the archery task than those in the control group. Additionally, beginner- and elite-level participants in the flotation-REST group experienced significantly lower average muscle tension during the archery task than participants of the same skill levels in the control group. However, no significant difference in performance during the archery task was observed between the flotation-REST and control group. The authors conclude that flotation-REST may assist athletic training by helping reduce physical stress and exertion.

Driller and Argus (2016) examined the effects of flotation-REST in 60 elite, international-level athletes across nine sports to determine if flotation-REST was a viable strategy for athlete recovery [40]. Following exercise training for their sport, each participant completed a 45-minute flotation-REST session. Pre- and post-float, participants completed the MDMQ as well as a questionnaire on perceived muscle soreness (MS). Participants also reported whether they had napped during the flotation-REST session or whether they had remained awake. The results found that, following the single flotation-REST session, participants reported a significant enhancement in 15 of the 16 MDMQ mood-state variables and significantly MS. Small to moderate effect sizes in favor of napping for 9 of the 16 MDMQ mood-state variables were also found when compared to the no-napping group, suggesting that napping in combination with flotation-REST may provide additional benefits for enhancing certain mood-state variables. In summary, participants saw an improvement in both mood state and muscle soreness, indicating that flotation-REST may be an effective tool for both physical and psychological recovery following training in elite athletes.

In 2018, Börjesson et al. sought to determine if flotation-REST might affect golfing performance [41]. 30 participants who were members of local golf clubs were recruited; each participant completed one 45-minute session of flotation-REST and one 45-minute control session (sitting quietly in an armchair), with sessions being separated by approximately one week. Following each session, heart rate and blood pressure were assessed, and participants completed the anxiety scales of the CSAI-2. Then, participants completed a golfing task while EMG was recorded to assess muscle tension. For the task, participants were required to execute straight golf putts towards a circular target at a distance of two, three, and four meters; this series was repeated four times for a total of twelve putts per participant. Performance on this task was measured via the final distance of the golf ball to the target. The results showed that there were no significant differences between the flotation-REST and control groups for blood pressure, heart rate, anxiety, muscle tension, or putting precision. Specifically, the flotation-REST and control sessions induced similar levels of reduction in blood pressure, heart rate, anxiety, and muscle tension. Thus, the authors conclude that flotation-REST may reduce stress and anxiety but may not directly improve golfing performance.

Broderick et al. (2019) examined the effects of flotation-REST on exercise recovery [42]. 19 male athletes completed a 1-hour flotation-REST session and a 1-hour passive recovery session in which they sat in a reclined chair in a dimly lit room. Before each treatment, participants completed an exercise task known as the Basketball Exercise Simulation Test. Upon completion of the test, post-exercise measures (performance tests, saliva collection, perceptual measures, and an algometer) were administered to determine the level of fatigue experienced by the participants. Once all post-exercise tests were completed, participants completed one of the recovery interventions. Participants then went home to sleep, during which their rest was monitored using a wrist actigraph, before returning the next morning to perform the same Basketball Exercise Simulation Test. Finally, post-exercise measures were recorded once more 24 hours after the first exercise task. The results showed that flotation-REST significantly enhanced measures of power and speed in participant’s performance the day following the treatment. Additionally, small to large effect sizes were found for all sleep measures (sleep quality, sleep latency, total sleep time, sleep efficiency, wake after sleep onset, awakenings per hour, and mean wake durations). The authors concluded that flotation-REST may serve as an effective tool for enhanced sleep quality and performance recovery in athletes.

#### 3.6.3. Physiological effects

We found eight studies that investigated the physiological effects of flotation-REST. Based on these studies, flotation-REST may induce physiological change. All eight studies found evidence in support of this conclusion, with two studies finding such evidence in regard to blood pressure (with one of these studies also finding evidence in regard to breathing rate and heart rate variability), two studies regarding cortisol, and one study each regarding right hemispheric processing, urinary VMA, EEG activity, and resting state functional connectivity. However, two studies found evidence suggesting that flotation-REST had no effect on luteinizing hormone; one of these studies also found no evidence of flotation-REST affecting cortisol, contradictory to the other two aforementioned studies, along with a similar lack of evidence for a host of other neuroendocrine measures [47]. Despite this, all studies (8 out of 8; 100.0%) found at least one significant result in favor of flotation-REST’s physiological effects. If we assume that the probability of a study finding a significant result is 50% (i.e., a coin toss), a Bayesian binomial test shows that there is very strong evidence (BF_+0_ = 56.778) in support of the hypothesis that the likelihood of a study finding that flotation-REST can produce physiological change is greater than chance. Given this and the high number of studies in this category relative to the other categories, we can reasonably assume that flotation-REST is capable of altering physiological functioning. We provide summaries for each of the eight studies below.

Fine and Turner (1982) conducted a pilot study in which they investigated the effects of flotation-REST on hypertension [43]. Three participants were involved in the study, identified as participants A, B and C. Participants completed 20 40-minute flotation-REST sessions over roughly a 10-week period, and blood pressure was taken before every flotation session. The STAI was also administered before and after each session to participant A. Post-treatment follow-ups were conducted in 3-month, 6-month, and 10-month increments. Flotation-REST resulted in significant reductions in both systolic and diastolic blood pressure after the treatment period that persisted throughout the follow-up sessions for all participants. Additionally, for participant A, anxiety tended to decrease from pre- to post-session, although whether this reduction was significant was not reported. The authors view their findings as tentative support for the use of flotation-REST as an intervention for reducing blood pressure.

In 1983, Turner and Fine investigated the effects of flotation-REST-induced relaxation on plasma cortisol, ACTH, and luteinizing hormone (LH) [44]. 12 healthy participants were randomly assigned to either a flotation-REST or a control group, in which participants rested in a reclining chair within a dimly lit room. Each group completed four 35-minute sessions, one a day for four consecutive days. Before the start of the treatment program, participants completed a baseline session during which blood samples were taken as participants quietly rested. During the treatment sessions, both groups underwent the same procedure meant to induce relaxation: first, the lights would dim until dark; then, a relaxing audio message would play for 90 seconds. Lights would then be brought back to the original dim lighting for 60 seconds before the cycle would repeat for a total of 10 times. Blood samples were drawn before and after the third treatment session and at a follow-up session four to five days after completion of the treatment program. At the end of the program, participants filled out a brief questionnaire surveying their overall relaxation experience. While both groups reported that their respective treatments were relaxing, only the flotation-REST group showed significant pre- to post-session changes in levels of cortisol and ACTH in the third treatment session. Additionally, cortisol levels during the follow-up session were significantly lower than cortisol levels during baseline for the flotation-REST group but not the control group. Conversely, no significant changes in ACTH or LH occurred between baseline and follow-up for either group. The authors suggest that flotation-REST may induce relaxation by decreasing activity in the pituitary-adrenal axis.

Turner and Fine (1991) examined the degree to which repeated flotation-REST sessions might lower plasma levels of cortisol as well as variability in plasma cortisol levels, effects that would promote an increased and stable state of relaxation [45]. 27 participants were recruited, pair-matched based on initial values of plasma cortisol, and then split into two groups: flotation-REST and control. Those in the control group rested in a fully reclined cushioned chair for their treatment sessions. All participants completed eight 40-minute sessions across 3 weeks. Beginning with the fifth session, blood samples were taken before and after each session. The results showed that both the average plasma cortisol concentration and variability significantly decreased across sessions in the flotation-REST group, whereas no significant change was detected in the non-REST group. These findings suggest that flotation-REST may induce a state of relaxation by reducing cortisol levels and cortisol level variability.

In 1994, Raab and Gruzelier sought to examine the effects of flotation-REST on right hemispheric processing [46]. 32 right-handed participants were recruited and randomly assigned to either the flotation-REST or the control group. Each participant began by completing HPT, in which letter and number objects were sorted by touch while blindfolded once, and the WRMT, in which participants were asked to identify visual stimuli in the form of words or faces they had previously seen. Then, those in the flotation-REST group completed a single 90-minute flotation session, while those in the control group were free to do as they pleased for the 90 minutes. Each participant was then re-administered the two tests. The results show that those in the flotation-REST group improved their performance on the HPT significantly more than those in the control group when the HPT was performed with the left hand, a function supported by the right hemisphere. Those in the flotation-REST group also improved their performance on the WRMT significantly more than those in the control group when the task assessed for memory only of faces, a function also supported by the right hemisphere. The authors conclude that flotation-REST enhances right hemispheric processing, and they note this enhancement was produced without any deleterious effects on left hemispheric processing, making flotation-REST preferrable to other forms of hemispheric processing enhancement such as hypnosis.

In the same year, Schulz and Kaspar sought to identify the neuroendocrine and psychological changes that occur during flotation-REST [47]. Five healthy males were recruited and completed one 60-minute flotation-REST session and one 60-minute control session, during which participants rested in a supine position in a dimly lit room. These sessions occurred at least one week apart, and the order of the sessions was randomized between-participants. Before the experiment, participants engaged in two habituation flotation-REST sessions over two to three weeks; two weeks prior to the start of the experiment, participants completed the MMPI [48] and SSS-IV [49]. 30 minutes before each session, blood samples were drawn, and additional samples were drawn every 15 minutes for 2 hours after the completion of the session. Before and after each session, participants completed the ZMQ [50]; participants also completed the SSS [51] and rated their levels of euphoria and relaxation using a VAS after each session. Biochemical analyses of the blood samples assessed the following: cortisol, TSH, prolactin, LH, GH, melatonin, ß-endorphin, ADH, T4, GABA, HVA, magnesium, urinary VMA, osmolality, and lactate. The results demonstrated no significant neuroendocrine changes attributable to flotation-REST across all participants, apart from a 37% average reduction in urinary VMA. Psychological results, however, indicated that sedation, euphoria, and relaxation were significantly greater after flotation-REST than the control treatment. The authors speculate that their negative neuroendocrine findings may potentially be due to the small sample size used in the study.

In 2001, Iwata et al. conducted additional analyses on the data collected in their 1999 study [52, 53]. EEG data was examined from two participants who had completed 1-hour flotation-REST sessions and had reported perceiving visual imagery while inside the float tank. Wavelet analyses of the EEG data revealed that the mean duration of theta activity was significantly longer than that of alpha activity. The authors conclude that occurrence of both theta and alpha activity and the difference in their durations accompanies the perception of visual hallucinations during flotation-REST.

In 2021, Al Zoubi et al. conducted the first functional neuroimaging study on flotation-REST to determine whether flotation-REST leads to altered patterns of resting-state functional connectivity (rsFC) [14]. 48 healthy participants were recruited and assigned to complete three 90-minute sessions of either flotation-REST or chair-REST, in which participants rested in a zero-gravity chair. Before and after the session, participants underwent functional magnetic resonance imaging (fMRI), during which they rested quietly. The authors then completed an exploratory analysis of the fMRI data using multivariate distance matrix regression (MDMR), a method used to find peak clusters of changes in connectivity between the pre- and post-float fMRI data. Results from this analysis showed significantly decreased rsFC within and between posterior hubs of the default-mode network as well as in cortical tissue in the somatomotor network, and a similar pattern of reduced rsFC was found in the post-chair REST fMRI data. Given that these brain networks are responsible for creating and mapping the sense of self, the authors suggest that flotation-REST may reduce self-reflective processes directed towards the body.

To explore the mechanism by which flotation-REST induces anxiolytic effects, Flux et al. (2022) investigated the acute cardiovascular effects of flotation-REST [54]. A total of 57 participants were recruited: 37 participants met the criteria for one or more anxiety disorders and thereby comprised the anxious group, while the remaining 20 participants did not meet the criteria and comprised the non-anxious group. All participants complete a single 90-minute flotation-REST and a control session that entailed watching a 90-minute nature film. The order in which participants completed these sessions was randomized, and sessions were spaced one week apart. Before and after each session, participants completed the STAI and the serenity scale from the PANAS-X. During each session, wireless and waterproof sensors attached to the participant measured heart rate, heart rate variability, breathing rate, and blood pressure. Relative to the film condition, for both participant groups, flotation-REST significantly lowered systolic and diastolic blood pressure, breathing rate, and certain metrics of HRV including the standard deviation of the interbeat interval, low frequency HRV, and very low-frequency HRV. There were no significant physiological differences between the anxious and non-anxious groups, except for a difference in breathing rate, where the anxious participants had a higher breathing rate in comparison to the non-anxious groups across both conditions. Blood pressure was the only physiological measure that significantly correlated with post-float reductions in state anxiety and increases in serenity. Based on these results, the authors suggest that flotation-REST may induce anxiolytic effects by lowering sympathetic arousal, thereby shifting the autonomic nervous system toward a more parasympathetic state.

#### 3.6.4. Stress

We found six studies that investigated the effects of flotation-REST on stress. Based on these studies, flotation-REST appears to be effective at reducing levels of stress. All six studies found evidence in support of this conclusion, with two studies finding such evidence in regard to clinical samples while the other four studies were in regard to non-clinical samples. Out of the four quantitative studies in this category, all studies (4 out of 4; 100.0%) found significant results in favor of flotation-REST reducing pain. If we assume that the probability of a study finding a significant result is 50% (i.e., a coin toss), a Bayesian binomial test shows that there is moderate evidence (BF_+0_ = 6.200) in support of the hypothesis that the likelihood of a study finding that flotation-REST can significantly reduce stress is greater than chance. Given this, flotation-REST may be a useful method for stress reduction. We provide summaries for each of the six studies below.

In 1983, Suedfeld et al. sought to resolve conflicting accounts of the effects of flotation-REST on stress [15]. 27 participants were and completed a single 1-hour long flotation-REST session. 5 measures were administered to each participant: the AST measure was administered pre-float; the SSS both pre- and post-float; and the BCS, RMS-person, and RMS-place post-float [55]. At the group level, there was a significant reduction in stress from pre- to post-float. Participants commonly reported feeling calm, still, and at rest and described the tank as a pleasant, relaxing experience. The authors suggest that flotation-REST generally reduces stress, and opposing accounts suggesting flotation-REST increases stress are likely attributable to the physical discomfort induced by earlier designs of the float pod.

In 1984, Jacobs et al. compared the effect of flotation-REST on relaxation to that elicited by a normal sensory environment [56]. 28 participants were recruited and randomly assigned to either a flotation-REST or a control group, in which participants relaxed in a supine position in a small room. Both groups completed 10 45-minute sessions; in each session, participants completed a simple relaxation program consisting of guided point-to-point relaxation, breathing techniques, and visual imagery techniques. Starting at the fifth session, physiological measures including electromyography (EMG), galvanic spin response (GSR), blood pressure, and skin temperature were taken pre- and post-session; participants also completed the SRQ post-session. From pre- to post-session, those in the flotation-REST group reported greater subjective relaxation and showed significantly greater reductions on systolic and diastolic blood pressure in comparison to the control group. Based on these findings, the authors suggest that flotation-REST, coupled with other relaxation techniques, may induce a deeper state of relaxation than either method alone.

Similarly, Forgays and Belinson (1986) evaluated the effects of flotation-REST on relaxation using both objective and subjective measures [57]. 30 participants were recruited; each participant completed three flotation-REST sessions that were a maximum of 150 minutes in length and spaced a week apart. During each session, heart rate was recorded; following each session, participants completed the SAS, MAACL, and ISQ, along with a brief interview about their experience in the float tank. The results showed that there was a significant effect of time on heart rate, such that heart rate dropped notably throughout most of the flotation-REST session before gradually increasing toward the end of the session. Additionally, during the interview, most participants reported that their experience in the float tank was positive and pleasant, and participants often underestimated the duration of their flotation-REST session. From these findings, the authors conclude that flotation-REST is a relaxing, pleasant experience.

In 1991, Forgays et al. compared the relaxation potential of standard flotation-REST with “dry” flotation-REST [58]. 24 participants were recruited; all participants completed a single 1-hour session of both standard “wet” flotation-REST and “dry” flotation-REST, in which participants laid supine on a waterbed mattress located in the same float tank. Sessions were spaced about one week apart. Before and after each session, participants completed the POMS and SPI; during each session, heart rate was recorded via waterproof electrodes. The results showed that there were no significant pre- to post-session changes in POMS and SPI scores across both groups. However, there was a significant difference in heart rate between the “wet” flotation-REST group and the “dry” flotation-REST group, such that those in the “wet” flotation-REST group experienced a greater decrease in heart rate from pre- to post-session. Additionally, there was a significant effect of gender on heart rate, such that females displayed a greater decrease in heart rate following both flotation-REST sessions compared to males. Based on the physiological results, the authors conclude that standard flotation-REST may induce greater levels of physical relaxation than “dry” flotation-REST.

In 2007, Asenlof et al. examined how flotation-REST might be used in tandem with psychotherapy to help alleviate severe stress caused by health problems [59]. Case studies were conducted on two women on long-term sick-leave, aged 55 and 58, and were carried out over a period of one year. One participant had been diagnosed with burnout depression while the other with fibromyalgia; both participants reported high levels of anxiety prior to the start of the treatment. Each participant completed a treatment program with several components: 35 sessions of flotation-REST every other week for 45 minutes per session and 8 sessions of group therapy, conversational therapy, and picture production (during which participants are asked to freely draw what comes to mind) each. Throughout the program, participants were asked to track their progress in journals and participate in two long-form interviews, one conducted halfway through the program and the other at the conclusion. The EPP method was then used to process the qualitative data. Both participants reported that they experienced positive changes to their physical, mental, and emotional well-being. Notably, negative feelings such as anger, frustration, grief, and fear subsided, and participants felt as if they had greater control over their life. The authors conclude that flotation-REST in combination with psychotherapy can holistically enhance well-being in individuals with high levels of stress.

To further explore the effects of combining flotation-REST with psychotherapy, Kjellgren et al. (2011) applied this treatment combination to six participants diagnosed with burn-out syndrome [10]. Participants completed a 10-week treatment program consisting of twice-weekly 45-minute flotation-REST sessions and weekly psychotherapy sessions. Participants were interviewed twice about their experiences with the treatment and its effects on their daily lives, with the first interview (about 30 minutes) conducted after four weeks from the start of the program and the last interview (about 60 minutes) conducted after the completion of the entire program. The EPP method was used to process the qualitative data, and results showed that the flotation-REST sessions induced deep relaxation and altered states of consciousness, with experiences like sensations of flying, entering a state of “nothingness”, and feelings of distinguishing the mind from bodily limitations commonly occurring. Additionally, some participants reported that the feeling that the mind and body were separate entities gave rise to personal insights concerning their ailments. Participants reported a heightened awareness of physical sensations, breathing patterns, bodily responses, body image, and body processes in general. By the end of treatment program, all the participants reported feeling “full of energy and strength” and were able to return to work full-time. Thus, the authors suggest that their findings serve as preliminary evidence in support of the use of flotation-REST and psychotherapy for chronic stress-related psychological issues.

#### 3.6.5. Consciousness

We found five studies that investigated the effects of flotation-REST on consciousness. Based on these studies, flotation-REST commonly appears to induce altered states of consciousness. All five studies found evidence in support of this conclusion, with participants across these studies reporting experiences of deep relaxation, visual and auditory hallucinations, and altered perceptions of time. Only one of these studies used quantitative methods for their main analyses; thus, there are not enough studies in this category alone to run statistical analyses. However, given the uniform agreement in the results of all five studies for the consciousness-altering effects of flotation-REST, we can reasonably assume that flotation-REST is capable of inducing these changes. We provide summaries for each of the five studies below.

In 1961, Lilly and Shirley investigated the “ego-altering” effects of flotation-REST [1]. A similar flotation environment to that of [60] was used, such that participants floated neutrally buoyant (via the use of weights) while upright and submerged in a tank inside of a stimulus-isolating room. Participants wore a stimulus-restricting mask that allowed for normal breathing while underwater while still limiting sensory input. An unspecified number of participants were recruited from the community. During the study, participants rotated among three different roles. Participants would first act as a regular participant floating within the tank and could set the duration of their flotation session. While floating, participants were asked to try to introspect and attend to their sensory experiences. Then, after exiting the tank, participants acted as “safety man” in which they would sit outside of the isolation area and observe another participant floating, operate the flotation equipment, and be on standby should any issues arise. Lastly, the participant became the “self-observer,” in which they were allowed to float without a safety man in the room, allowing for maximum isolation and ego freedom. Participants’ anecdotal reports showed that many participants experienced altered states of consciousness, such that visual and auditory hallucinations, perceptions of body distortions, and mood swings were frequently experienced. Notably, a number of participants experienced negative effects due to physical discomfort or fear as a result of the flotation procedure. The authors suggest that positive flotation experiences may occur only when internal and external stimuli, particularly any discomforts, are minimized.

Iwata et al. (1999) sought to investigate the consciousness-altering effects of flotation-REST through the lens of sleep physiology [52]. Five participants were recruited; each participant completed two 1-hour flotation-REST sessions while a polysomnograph was recorded. After each session, participants were interviewed about their subjective experiences while inside the float tank. The results indicated that participants’ experiences could be classified into two categories: participants either reported perceiving visual images which were accompanied by alpha and theta waves as well as frequent rapid eye movements (REM), or participants fell into a deep sleep without perceiving any visual imagery. Participants who did perceive visual imagery noted that these images were more vivid or clear than those perceived during typical dreaming. The authors suggest that flotation-REST may induce altered states of consciousness that are different from the conventional states of wakefulness and sleep.

Norlander et al. (2000) examined whether an individual’s experience during flotation-REST might be modulated by that individual’s previously experienced altered states of consciousness or the contextual setting in which the flotation takes place [61]. Two groups of participants were recruited: a group of 14 participants who were former drug-users, and a group of 14 participants of comparable age, gender, and occupation who had no drug experience. Participants in these groups were then randomly assigned to one of two experimental conditions: a “strict” condition, in which participants completed the study in a standard flotarium setting, and a “fantasy” condition, in which the flotarium was decorated to be vividly colored, and participants listened to stories of the fantastical experiences reported by previous float tank users before their own session. All participants completed a single 1-hour flotation-REST session. Pre- and post-float, participants completed a questionnaire assessing pain and mood. Additionally, the post-float questionnaire included questions evaluating the flotation experience. The results showed that there were no significant differences in flotation-REST effects between the two groups or the two conditions, suggesting that earlier experiences of altered states of consciousness as well as contextual setting do not affect flotation-REST. Additionally, there were significant changes in pain and mood, such that participants experienced reduced pain and enhanced mood from pre- to post-float. Participants reported a variety of experiences stemming from the altered state of consciousness induced by flotation-REST, including visual and auditory hallucinations, altered time perception, a sense of weightlessness, out-of-body experiences, a sense of cosmic unity, and visions of one’s own birth. From these results, the authors conclude that flotation-REST stands as a promising method for inducing altered states of consciousness.

In 2004, Kjellgren et al. investigated the relationship between the degree of altered state of consciousness induced by flotation-REST and subjective pain and stress thresholds [12]. 23 athletes were recruited and completed one 45-minute flotation-REST session and one 45-minute chamber-REST session six weeks apart, and the order of which was randomized between-participants. Immediately after each REST session, participants’ degree of altered consciousness was measured using the EDN questionnaire. Then, participants completed an experimental pain procedure, which involved tightly binding one of the participant’s arms with a rubber band. During this procedure, pulse rate and oxygen levels were measured using a pulse oximeter, and subjective levels of pain and stress were reported at minute increments throughout. The pain procedure lasted no more than 15 minutes, and participants were asked to estimate the duration of the procedure after its completion. Results showed that flotation-REST induced a significantly higher degree of altered state of consciousness compared to chamber-REST. Additionally, participants who reported a higher degree of altered state of consciousness during flotation-REST experienced significantly higher levels of subjective pain and stress during the pain procedure while perceiving the procedure duration to be shorter than participants with a lower degree of altered state of consciousness during flotation-REST. Thus, the authors suggest that pain and stress experienced during higher degrees of altered state of consciousness may be perceived as more intense and acute, perhaps due to increased sensitivity to sensory stimuli.

In 2008, Kjellgren et al. sought to qualitatively assess the general experiences of flotation-REST among experienced users [62]. Eight participants were recruited, each of whom was diagnosed with depression, burn-out syndrome, or chronic pain; had participated in flotation-REST at least eight times; and who had completed their most recent flotation session within the last month. Participants completed a 45–60-minute interview, during which they were asked to reflect on their experiences during flotation-REST and the impact of flotation-REST on their life. The EPP method was used to analyze the interview transcripts. Overall, participants reported flotation-REST to be a pleasant experience that often produced pain relief. Participants reported frequently experiencing altered states of consciousness during flotation-REST, varying from milder states involving deep relaxation and altered time perception to more powerful states with notable perceptual changes such as out-of-body experiences and perinatal experiences. The authors conclude that, while flotation-REST may induce powerful mental experiences, these experiences are typically positive and productive.

#### 3.6.6. Psychology

We found five studies that investigated the effects of flotation-REST on psychology. Based on these studies, flotation-REST may induce positive psychological change. All five studies found evidence in support of this conclusion, with all of the studies finding evidence of reduced anxiety, three of decreased depression, and two of decreased arousal. Out of the four quantitative studies in this category, all studies (4 out of 4; 100.0%) found significant results in favor of flotation-REST reducing pain. If we assume that the probability of a study finding a significant result is 50% (i.e., a coin toss), a Bayesian binomial test shows that there is moderate evidence (BF_+0_ = 6.200) in support of the hypothesis that the likelihood of a study finding that flotation-REST can significantly alter one’s psychological state is greater than chance. Overall, flotation-REST appears to lead to improvements in psychological well-being. We provide summaries for each of the five studies below.

Forgays and Forgays (1994) sought to determine the effects of flotation-REST on mood in those with Type A personalities, which are characterized by impatience, aggression, and competitiveness [63]. 40 participants were recruited and were separated into either a Type A group or Type B group, depending on their scores on a personality assessment. Participants were then randomly assigned to complete a single 1-hour session of either flotation-REST or a control session, in which they rested quietly in a room. Before and after each session, participants completed the SPI and POMS measures. The results showed that levels of anxiety, anger, depression, confusion, vigor, hostility, and fatigue all significantly decreased from pre- to post-float. Additionally, these decreases were found in comparable levels across all participants, such that there was no significant effect of Type A/Type B personality on pre- to post-float scores. Thus, the authors conclude that flotation-REST may help reduce certain Type A characteristics, regardless of what type of personality that individual originally displays.

In 1995, Suedfeld and Eich conducted a two-part study in which they examined the effect of flotation-REST on mood and autobiographical memory [64]. For both parts of the study, the POMS questionnaire, which assesses levels of anxiety and arousal, was administered before and after each treatment session. The first study recruited 32 participants and randomly assigned them to one of two treatment groups: the flotation-REST group, in which participants rested in a supine position within a float tank for 1 hour; or the control group, in which participants simply waited in the university’s psychology building and returned to the laboratory after 1 hour. The results showed that the flotation-REST group scored significantly lower than the control group on both the POMS anxiety and arousal scales post-session. The second study recruited 24 participants who were also assigned into flotation-REST and control groups. All participants began by completing two measures assessing their current levels of mood and arousal. Then, during their assigned treatment, participants were asked to complete the same measures, either over an intercom system for participants in the flotation-REST group or on pen-and-paper for those in the control group. Subsequently, participants completed a new task in which they had 120 seconds to retrieve a specific autobiographical memory in response to a list of neutral probe words. Participants were then asked to date their retrieved event and describe the event using several rating scales. Participants in the flotation-REST group in the second study experienced comparable changes in anxiety and arousal to participants in the flotation-REST group in the first study. Additionally, participants who completed flotation-REST in the second study reported that their autobiographical memories were significantly more pleasant, vivid, and emotionally intense than participants in the control group. Thus, the authors conclude that flotation-REST may induce changes in mood and arousal that then affect the perception of autobiographical memories.

Kjellgren et al. (2013) reported the subjective experiences of an individual who had completed two and a half years of flotation-REST (totaling 75 sessions) in order to treat a number of neuropsychiatric and mental health disorders [65]. The participant was a 24-year-old woman diagnosed with attention deficit hyperactivity disorder, atypical autism, post-traumatic stress disorder, anxiety, and depression. The participant completed a semi-structured interview about her experiences with flotation-REST and its impact on her well-being. The EPP method was used to analyze the interview transcript. The participant reported experiencing an overall improved quality of life, crediting her enhanced well-being and healthier behavior to flotation-REST. The participant also reported no negative effects from her treatment. From this case study, the authors conclude that flotation-REST may have therapeutic effects on a wide range of mental health issues.

In 2014, Kjellgren and Westman evaluated the psychological effects of flotation-REST in healthy participants [13]. Based on previous studies, the authors hypothesized that flotation-REST would have beneficial effects on levels of pain, depression, anxiety, stress, energy, optimism and sleep quality. 65 participants were recruited and were randomly assigned into a flotation-REST group or a wait-list control group. Those in the flotation-REST group completed 12 45-minute sessions across a 7-week period. These participants completed a questionnaire containing the following measures: SE, HADS, LOT, SQ, MAAS, EDN, and two VAS assessing normal pain and experienced worst pain. Those in the control group completed the same questionnaire before and after a 7-week period. The results showed significant decreases in experienced stress, pain, anxiety, and depression as well as significant increases in sleep quality and optimism for the flotation-REST group compared to the control group. Additionally, there was a significant correlation between mindfulness in daily life and the degree of altered state of consciousness experienced during flotation-REST. The authors suggest that flotation-REST produces beneficial effects on mental health, even for healthy individuals.

In 2020, Khalsa et al. investigated the safety and tolerability of flotation-REST for individuals with anorexia nervosa [66]. Twenty-one partially weight-restored participants with anorexia nervosa completed a protocol involving four sequential sessions of REST: reclining in a zero-gravity chair (chair-REST), floating in an open pool (open flotation-REST), and two sessions of floating in an enclosed pool (closed flotation-REST). All sessions were 90 minutes and spaced approximately one week apart. Orthostatic BP was measured before and immediately after each session, in addition to measurements collected every 10 min during the session using a wireless waterproof system. Each participant’s affective state, awareness of interoceptive sensations, and body image were assessed before and after every session using the STAI, PANAS-X, IAM, BAS-2, BISS, PFRS, and EDEQ-6. The results indicated that there was no evidence of orthostatic hypotension or adverse events, such as panic attacks, severe dysphoria, or suicidal ideation, following all types of REST. All types of REST induced statistically significant reductions in blood pressure, anxiety, and negative affect; heightened awareness of cardiorespiratory sensations; and reduced body image dissatisfaction. These findings suggest that individuals with anorexia nervosa not only can safely tolerate the effects of flotation-REST but may also experience benefits relative to both their condition and their general mental well-being.

#### 3.6.7. Creativity

We found five studies that investigated the effects of flotation-REST on creativity. Based on these studies, flotation-REST appears to generally enhance creativity. One study found evidence that flotation-REST improves scientific creativity; one study found evidence that flotation-REST increases divergent thinking; one study found evidence that flotation-REST improves technical musical ability; and two studies found evidence that flotation-REST increases originality, although one of these studies also found that flotation-REST may impair creative problem-solving. Overall, however, all five studies (5 out of 5; 100.0%) found at least some significant result in favor of the creativity-enhancing effects of flotation-REST. If we assume that the probability of a study finding a significant result is 50% (i.e., a coin toss), a Bayesian binomial test shows that there is strong evidence (BF_+0_ = 10.500) in support of the hypothesis that the likelihood of a study finding that flotation-REST can significantly improve creativity is greater than chance. In conclusion, flotation-REST may enhance certain types of creative behavior, particularly those that rely on original thinking, while impairing other types of creative behavior, such as those that rely on problem-solving. We provide summaries for each of the five studies below.

Suedfeld and Baker-Brown (1987) sought to examine the effects of flotation-REST on scientific creativity [67]. Five full-time faculty members from the Department of Psychology at the University of British Columbia were recruited. Each participant completed six 1-hour flotation-REST sessions and six 1-hour sessions of sitting alone in their office in a counter-balanced order. During the office sessions as well as for the 30-minute period after their flotation-REST sessions, participants dictated ideas related to their scientific work into a tape recorder. Prior to the start of their dictation, participants also completed the POMS. Several months after their sessions, participants received transcripts of their tape recordings and were asked to note the following about their dictation: (1) the number of times participants changed from one idea to the next; (2) the number of times a novel idea came to mind; (3) the level of creativeness of each idea rated on a scale from 1-10; and (4) the number of ideas that eventually led to new research publications, grant proposals, or similar accomplishments. The results indicated that ideas generated after flotation-REST were rated as significantly more creative than those generated during office settings. Additionally, flotation-REST was associated with lower levels of negative mood. The authors conclude that flotation-REST may facilitate creative behavior along with positive affect.

Forgays and Forgays (1992) evaluated the effects of flotation-REST on divergent thinking [5]. 30 participants were recruited and assigned to complete a single 1-hour session of either flotation-REST or relaxing in a dimly lit room (control group). All participants completed the GCS before and after the session as well as the SPI and POMS, which were used to assess affect. Those in the flotation-REST group showed significant increases on GCS scores pre- to post-float while those in control group did not. Additionally, those in the flotation-REST group reported decreases in anxiety/tension, depression, hostility, and fatigue along with increases in vigor. The authors speculate that these changes in mood may contribute to the observed enhancement in creativity.

In 1998, Norlander et al. conducted two studies to investigate the effects of flotation-REST on creative problem-solving and originality [68]. In the first study, 40 participants were recruited and randomly assigned to either a flotation-REST group or a control group, in which participants sat in an armchair. Prior to the session, participants completed the FS test. Then, participants worked on a creative problem-solving task called Silveira’s cheap necklace problem [69] for 5 minutes before completing their assigned treatment for 45 minutes. After the 45 minutes had elapsed, participants continued to work on the problem until either completion or after 25 minutes had elapsed. The results showed that the flotation-REST group took a significantly greater amount of time to solve the problem compared to the control group; the authors suggest that this indicates that flotation-REST may impair creative problem-solving. In the second study, 54 participants were recruited and randomly assigned to one of three conditions: flotation-REST, chamber-REST, or non-REST, in which participants sat in an armchair. Prior to the session, participants completed the FS test. Then, participants completed their assigned treatment for 45 minutes. Following the treatment, participants completed the Syllogisms I and FREGO tests, along with a novel task designed to assess originality. The results showed that the flotation-REST group scored significantly higher on originality in comparison to the chamber- and non-REST groups. The authors conclude that flotation-REST may increase original thinking while impairing creative problem-solving abilities.

Norlander et al. (2003) conducted two additional studies to examine the effects of flotation-REST on divergent creativity, logical thinking, and essay-writing [70]. In the first study, 38 participants were recruited and randomly assigned to complete either a single 45-minute flotation-REST session (single-flotation group) or three 45-minute flotation-REST sessions (three-flotations group). Before either their single session (for the single-flotation group) or their third session (for the three-flotations group), participants completed the LOT, Syllogisms test (either version I or II), and beer can/brick test (either beer can or brick). After their session, participants completed the Syllogisms and beer can/brick tests again but were administered the versions that they had not received pre-float. The results showed that there were no significant differences in divergent creativity or logical thinking between the two groups, such that improvements in creativity ability and reductions in logical reasoning were comparable. In the second study, a 2×2 design was used: 32 participants were recruited and randomly assigned to either a flotation-REST or chamber-REST group and a stress or non-stress group. Participants completed the FS, LOT, and HADS measures before their session and the EDN measure after their session. Prior to their treatment session, those in the stress group also completed an additional task meant to induce stress by pressuring participants to complete a series of tests given unclear instructions and a time limit. Participants then underwent their assigned REST for 45 minutes. Following each session, participants were tasked with writing an essay which was later assessed for elaboration, liveliness, originality, and realism. Analysis of the essays revealed that the flotation-REST group showed significantly greater originality while the chamber-REST group showed significantly more elaboration and realism. Additionally, for those in the chamber-REST group, participants who completed the stress task showed significantly greater realism in their essays compared to those that did not. Finally, because the flotation-REST group experienced a significantly greater degree of altered state of consciousness than the chamber-REST group, the authors suggest that a greater degree of altered consciousness may promote original thinking whereas a lower degree of altered consciousness may support more realistic thinking.

Vartanian and Suedfeld (2011) examined the effects of flotation-REST on jazz improvisation [71]. 13 participants who were enrolled in an intermediate-level jazz improvisation college course were recruited. Participants were randomly assigned to a flotation-REST group, in which four 1-hour sessions were completed across four weeks, or a control group. All participants completed a recorded 5-minute improvisational performance both one week before treatment and one week after treatment (or, for the control group, before and after the same four-week period). Each recording was then evaluated by the jazz improvisation class instructor, who was blinded to each participant’s assigned condition, on five dimensions: improvisation, creativity, expressiveness, technical ability, and overall quality. Following the four-week period, the instructor was also asked to evaluate each participant’s change in improvisational ability on the same five dimensions based on their course performance. The results showed that the flotation-REST group scored significantly higher on technical ability based on both their post-treatment improvisation recordings and instructor’s course evaluations compared to the control group. No other significant differences between the two groups were found, other than that the flotation-REST group had a significantly higher final class grade on average. The authors suggest that flotation-REST may improve perceptual-motor skills involved in jazz improvisation.

#### 3.6.8. Clinical anxiety

We found four studies that investigated the effects of flotation-REST on clinical anxiety. Based on these studies, flotation-REST appears to be an effective therapeutic intervention for a range of clinical anxiety disorders. All four studies found evidence in support of this conclusion, with two studies finding such evidence specifically in regard to GAD and the other two regarding a variety of anxiety-related disorders. Only three studies in this category used quantitative methods for their main analyses; thus, there are not enough studies in this category alone to run statistical analyses. Overall, flotation-REST may be a promising treatment for clinical anxiety. We provide summaries for each of the four studies below. We provide summaries for each of the four studies below.

In 2016, Jonsson and Kjellgren evaluated the effects of flotation-REST on general anxiety disorder (GAD) [6]. 50 participants who met the criteria for GAD based on their responses to the GAD-Q-IV were recruited. Participants were randomly assigned to either a flotation-REST group or a wait-list control group. Those in the flotation-REST group completed a 7-week treatment program involving 12 45-minute flotation-REST sessions. All participants completed assessments at baseline, four weeks into the treatment program (or corresponding time for those in the control group), and at the end of the 7 weeks, as well as at a six-month follow-up. These assessments included the GAD-Q-IV, MADRS-S, PSQI, DERS, MAAS, and EDN scale. The results showed that those in the flotation-REST group experienced a significant reduction in GAD-symptomatology, whereas those in the control group did not. Additionally, flotation-REST was found to significantly improve sleep quality, emotional regulation, and depression symptoms. Aside from depression, these positive benefits were maintained at the 6-month follow-up. The authors suggest that flotation-REST may act as an effective complementary treatment to existing interventions for GAD.

In the following year, Jonsson and Kjellgren presented their findings from semi-structured interviews conducted with nine of the GAD-diagnosed participants in their 2016 study [7]. These participants had been assigned to the flotation-REST group and completed the interviews, which lasted from 45 to 75 minutes, after completing the treatment program. The EPP method was used to analyze the interview transcripts. While participants reported experiencing initial difficulties with achieving a state of relaxation in the float tank, they came to view the float tank as a safe, secluded, relaxing environment over the course of the program. During flotation-REST sessions, participants reported experiencing altered states of consciousness marked by changes in body perception, visual hallucinations, and distorted time perception. Throughout the treatment program, participants felt as if they gained greater levels of self-understanding and self-acceptance which led to improvements in mood, the adaptation of more optimistic attitudes, and better coping strategies, ultimately enhancing their quality of life. The authors suggest that flotation-REST may help individuals with GAD address the core issues underlying their condition, thereby resulting in comprehensive positive change.

Feinstein et al. (2018) examined the effects of flotation-REST on symptoms of anxiety, stress, and depression in a sample of 50 participants diagnosed with either depression, anxiety, a stress-related disorder (e.g., PTSD, GAD, panic disorder), or a combination thereof [72]. Participants a single one-hour session of flotation-REST, and a questionnaire containing the STAI, PANAS-X, KSS, WBPS, and VAS measures for relaxation, muscle tension, stress, depression, contentedness, refreshment, and energy were completed 30 minutes before and after the session. The results demonstrated that a single one-hour session of flotation-REST caused a reduction in anxiety across all participants, such that there was a significant decrease in anxiety at the group level. Flotation-REST also led to a significant decrease in stress, muscle tension, pain, depression and negative affect and a significant increase in reported feelings of serenity, relaxation, happiness, positive affect, overall well-being, energy, contentedness, and refreshment. Notably, the most severely anxious participants reported the largest changes in scores. These findings suggest that flotation-REST may be a promising technique for acutely reducing symptoms of anxiety and depression, although the authors note that the persistence of these effects is unclear.

To further investigate the anxiolytic effects of flotation-REST, in the same year, Feinstein et al. assessed the effects of flotation-REST on individuals with high anxiety sensitivity, or the fear of experiencing anxiety-related bodily sensations [73]. 31 participants, all diagnosed with either anxiety or depression, were randomly assigned to complete either a 90-minute session of flotation-REST or to watch a 90-minute nature documentary from the BBC Planet Earth series (representing the control group). All participants completed the PANAS-X, STAI, and VAS measures on relaxation and muscle tension 30 minutes before and after their treatment session. Those in the flotation-REST group experienced a significantly greater reduction in anxiety, muscle tension, and blood pressure as well as a significantly greater increase in levels of relaxation and serenity compared to the control group. Notably, those in the flotation-REST group also reported experiencing significantly enhanced interoceptive awareness and attention to cardiorespiratory sensations. The authors speculate that flotation-REST may reduce anxiety by facilitating a state of mindfulness via focused attention on current bodily sensations.

#### 3.6.9. Sleep

We found three studies that investigated the effects of flotation-REST on sleep. Based on these studies, flotation-REST may have limited benefits for individuals with sleep-related disorders. Two studies found evidence of flotation-REST improving sleep latency, but one of these studies found that this improvement only reached significant levels three months after treatment completion, while the other study found that only half of the participants experienced improvement. Additionally, the former study found comparable improvements via relaxation exercises. Regardless, flotation-REST appears to induce an acute state of restfulness similar to that experienced during stage I sleep. Unfortunately, there are not enough studies in this category alone to run statistical analyses. In conclusion, flotation-REST may not be particularly effective for sleep-related disorders, although it may still benefit non-clinical populations [17]. We provide summaries for each of the three studies below.

In 1989, Ballard evaluated the long-term effectiveness of flotation-REST on persistent insomnia [74]. 36 participants diagnosed with persistent insomnia were recruited and randomly assigned to one of four conditions: flotation-REST, relaxation exercises, flotation-REST coupled with relaxation exercises, or a waitlist control. Participants in one of the three active treatment groups completed four 45-minute treatment sessions over two weeks. Both subjective and objective measures were used to assess sleep latency at baseline as well as one, four, and twelve weeks after treatment. Namely, participants reported their sleep latency in a daily sleep log, while participants also used a Somtrak sleep assessment device to collect objective latency data. Participants also completed the BDI, POMS, and SRAS at baseline and each follow-up session. The results showed that flotation-REST, both by itself and in combination with relaxation exercises, led to significant reductions in subjective and objective sleep latency from baseline to the 12-week follow-up session. Notably, however, the immediate and short-term effects of flotation-REST on sleep latency were nonsignificant, such that the reductions in sleep latency decreased over time but did not reach a significant level until twelve weeks after the end of the treatment. Additionally, a comparable reduction in sleep latency was obtained through relaxation exercises only, such that there was no significant difference between reductions in sleep latency between the three active treatment groups. Despite these caveats, the author concludes that flotation-REST can improve sleep latency for those with persistent insomnia.

In 2017, Dunham et al. sought to compare the Bispectral Index (BIS) values obtained during flotation-REST and those found during sleep [75]. BIS monitoring is a method in which raw EEG signals are analyzed to assess the level of consciousness of an individual. Dunham acted as the sole participant in the study and underwent 22 1-hour flotation-REST sessions over the course of four months. During sessions 14 and 16, BIS values were recorded every minute. Additionally, the participant assessed their mood state (namely, their levels of relaxation and freshness) before and after each session. The results indicated the BIS values associated with flotation-REST were comparable to literature-derived BIS values during stage I sleep, such that the mean percent difference between the BIS values from the two flotation-REST sessions and the BIS values from stage I sleep were relatively low compared to the BIS values from a state of alertness or other stages of sleep. The authors conclude that flotation-REST may induce a state of rest analogous to that during stage I sleep.

Norell-Clarke et al. (2022) sought to assess the potential of flotation-REST as a treatment for insomnia [76]. 6 participants who had been diagnosed with insomnia were recruited, and all participants completed 12 45-minute sessions of flotation-REST over a 7-week period. Participants recorded sleep onset latency, wake after sleep onset, total sleep time, and sleep efficiency in a daily sleep log. Additionally, participants completed the ISI and MADRS-S to assess insomnia and depressive severity, respectively, at baseline, after treatment, and at a 2-month follow-up. The results were mixed; some participants did not report any beneficial effects from flotation-REST, while three participants reported improvements in sleep onset latency or wake after sleep onset. Additionally, two participants improved on sleep efficiency. Whether these changes were significant was not reported. Because the two participants who experienced the most improvements in their insomnia symptoms were in their 20s, the authors suggest that flotation-REST may be most beneficial as an insomnia treatment for young adults.

#### 3.6.10. Smoking cessation

We found two studies that investigated the effects of flotation-REST on smoking cessation. Based on these studies, flotation-REST may not be particularly effective as a therapeutic intervention for smoking cessation. Both studies in this category found evidence of other interventions, such as chamber-REST, being more effective at helping individuals reduce their levels of smoking than flotation-REST. Unfortunately, there are not enough studies in this category alone to run statistical analyses. In conclusion, flotation-REST may not facilitate decreased smoking behavior any better than other established interventions. We provide summaries for the two studies below.

In 1987, Suedfeld and Baker-Brown evaluated the effects of flotation-REST on smoking cessation in comparison to various forms of chamber-REST [77]. Namely, a 2×4 design was used for the chamber-REST component of the study: session duration (12 versus 24 hours) x presentation schedule of motivation messages designed to encourage the participant to quit smoking (all at once, distributed, self-paced, or no messages). 83 participants who smoked at least one pack of cigarettes daily for five years were recruited and randomly assigned to either one of the eight chamber-REST conditions or the flotation-REST condition, which involved five 1-hour flotation-REST sessions that also included the presentation of motivational messages. Participants were interviewed at a 3-month and 12-month follow-up about their smoking habits. The results showed that most of the chamber-REST groups experienced significant reductions in mean smoking (i.e., number of cigarettes per day) that persisted at both the 3-month and 12-month follow-up. Additionally, there were no significant differences resulting from the session duration or message presentation schedule. While flotation-REST also led to a significant reduction in mean smoking at the 3-month follow-up, this effect did not persist at the 12-month follow-up. Thus, the authors recommend chamber-REST above flotation-REST as a therapeutic intervention for smoking cessation.

In the same year, Forgays sought to determine the effectiveness of flotation-REST as a therapeutic intervention for smoking cessation [78]. 33 participants were recruited; participants smoked an average of 30 cigarettes per day for 20 years, and all participants reported having made a previous effort to stop smoking. A 2x 3 design was used: participants either completed four 60-hour flotation-REST sessions on consecutive days, five 150-minute flotation-REST sessions spaced a week apart, or no flotation-REST sessions (control); additionally, participants either did or did not receive motivational messages designed to encourage smoking cessation while in the float tank or via phone call (for the control group). Participants reported their smoking behavior (i.e., number of cigarettes per day) after completion of the treatment program as well as at a 12-month follow-up. The results showed that all groups displayed a large decrease in mean smoking following the treatment program, although whether these decreases were significant was not reported. No effect of motivational messages on smoking behavior was found. Those in the 150-minute flotation-REST group demonstrated a greater reduction in smoking at the 12-month follow-up in comparison to those in the 60-minute flotation group. Surprisingly, however, those in the control group demonstrated the greatest reduction in smoking at the 12-month follow-up compared to both flotation-REST groups. Thus, flotation-REST may not be particularly effective for smoking cessation, especially when compared to other interventional techniques.

#### 3.6.11. Other

In 1960, Jay Shirley sought to refine the procedural methods used in flotation-REST [60]. Specifically, Shirley wanted to standardize what he considered the three fundamental aspects of flotation-REST: the physical aspect, physiological aspect, and psycho-social aspect. For the physical aspect, Shirley noted that it was important to create an environment that obtained the maximum achievable reduction of ambient physical stimuli along with a dynamic maintenance of ambient temperature. To achieve these conditions, Shirley built a two-room laboratory, in which the float tank was housed, that substantially reduced incoming light, sound, vibration, and odor. Next, for the physiological aspect, Shirley sought to eliminate all sources of pain and discomfort stemming from body position, reduced blood flow, or abdominal distention. Thus, a float tank was designed that allowed participants to float upright while comfortably wearing both a stimulus-restricting oxygen mask and body weights that maintained neutral buoyancy and full immersion. Lastly, for the psycho-social aspect, Shirley speculated that certain types of persons were optimal participants for float-REST studies. Such participants were skilled in self-observation, memory, and attention to detail and were able to communicate their experience fully and freely with minimal distortion. Based on these criteria, Shirley recruited twelve participants, and each participant was free to choose the length of their flotation session, with the longest session lasting for four hours. In the form of anecdotal notes, participants reported experiencing emotions that shifted randomly from calm, contemplation, anxiety, elation, and depression. Some participants also experienced visual and auditory hallucinations along with hyperawareness of their bodily functions, such as being able to hear the sounds of their heartbeat. Overall, this study helped define the optimal conditions for flotation-REST and laid the groundwork for the float tank to be used as a tool for probing a wide range of psychophysiological phenomena.

In 1989, Turner et al. investigated whether light exposure might modulate the effects of flotation-REST [79]. 21 participants were recruited and randomly assigned into one of two groups: flotation-REST in the presence of light (REST-L) or flotation-REST in the absence of light (REST-D). All participants completed eight 40-minute flotation-REST sessions across four weeks. Eight blood samples were taken before the start of the flotation-REST program, and eight samples were taken during the last two weeks of the flotation-REST program, for a total of 16 blood samples per participant. Blood pressure measurements were also taken before and after each blood draw. At both baseline and after the completion of the program, participants completed the POMS scale. The results indicated that there were no significant differences between the REST-L and REST-D groups, such that groups experienced comparable improvements in mood, decreases in plasma cortisol, and decreases in mean arterial pressure. Thus, the authors conclude that exposure to light during flotation-REST does not compromise the benefits of flotation-REST.

Suedfeld et al. (1994) conducted four studies to test a hypothesis aiming to explain the wide-ranging effects of flotation-REST [80]. The Dynamic Hemispheric Asymmetry (DHA) model proposes that flotation-REST increases activity in the side of the cortical hemisphere that tends to be less dominant (typically thought to be the left hemisphere in most people), and this increased activation is responsible for the perceptual and mood changes accompanied by flotation-REST. Under the model, the authors posit that flotation-REST should lead to the following four changes: 1) increased perceptual recognition of objects based on visually incomplete representations, 2) improved story-telling abilities (i.e., more creative, complex stories), 3) decreased asymmetry of finger-tapping speed between the dominant and non-dominant hands, and 4) increased EEG activity in the non-dominant brain hemisphere. Four studies were conducted to test each of the hypotheses; 25, 19, 25, and 10 participants were recruited for each study, respectively. For participants assigned to the flotation-REST group in each study, a single flotation-REST session that was either one hour long (studies 1, 2, and 4) or two hours long (study 3) was completed. Based on the results of each study, evidence was found in support of hypothesis 3, whereas contradictory evidence was found for hypotheses 1 and 4, and evidence for hypothesis 2 was inconclusive. Based on the mixed results, the authors conclude that the DHA model fails to adequately explain the effects of flotation-REST

Sakata et al. (1995) tested the hypothesis that flotation-REST would enhance an individual’s ability to generate random sequences [81]. 7 participants were recruited, and all participants completed two types of treatment sessions in a counterbalanced order: 1) flotation-REST), and 2) bed-REST, in which participants rested in a supine position on a bed within a dark, quiet room. Participants completed in a total of two float-REST sessions and one bed-REST session with each session lasting 40 minutes. Additionally, there was a one-week interval between each float-REST session and a month-long interval between the two treatment types. Prior to each treatment session, electrodes were attached to the participant in order to measure EEG, EOG, heart rate, and respiration. Before each session, participants completed a task in which they were asked to orally generate a list of numbers while attempting to be as random as possible. Participants completed this same task immediately after the end of the treatment session as well as 40 minutes after the REST session. Results indicated that randomization indices scores were lower (indicated greater randomness) immediately after flotation-REST and that these low scores persisted even at the 40-minute post-session mark. Conversely, randomization indices scores increased after the bed-REST and persisted into the post-REST mark. Thus, the authors conclude that random number generation was significantly enhanced by flotation-REST in comparison to bed-REST, and they hypothesize that the deep relaxation and hypnagogic state induced by flotation-REST is what facilitates enhanced randomness.

## 4. Bias Assessment

In our paper, we systematically assess several categories of bias for each study under review. Firstly, we will evaluate the presence of selection bias, which pertains to potential systematic differences between participants selected for the study and those not included. This assessment will involve scrutinizing the methods used for participant recruitment and selection, as well as the adequacy of randomization procedures, to ensure the sample’s representativeness. Secondly, we will consider performance bias, which addresses variations in the administration of interventions or treatments between groups. This analysis will involve examining the standardization and consistency of procedures across different study arms. Thirdly, we will assess detection bias, focusing on the consistency and blinding of outcome assessments to prevent biased measurement or interpretation of results. Additionally, we will explore attrition bias, which involves examining the differential dropout rates between study groups and the potential impact on outcome data completeness and validity. Finally, we will examine reporting bias, which concerns the selective reporting of outcomes and results. Our thorough evaluation of these bias categories will ensure a comprehensive understanding of the methodological rigor and validity of the studies under review in our paper. Table 4 summarize the bias assessment for each study based on selection, performance, detection, attrition, and reporting bias.

**Table 4.**
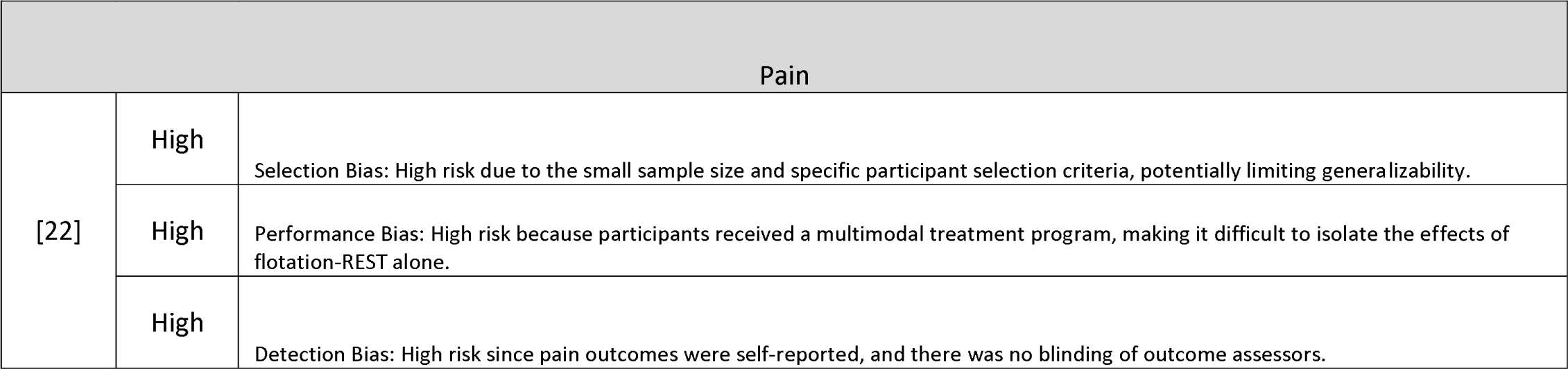
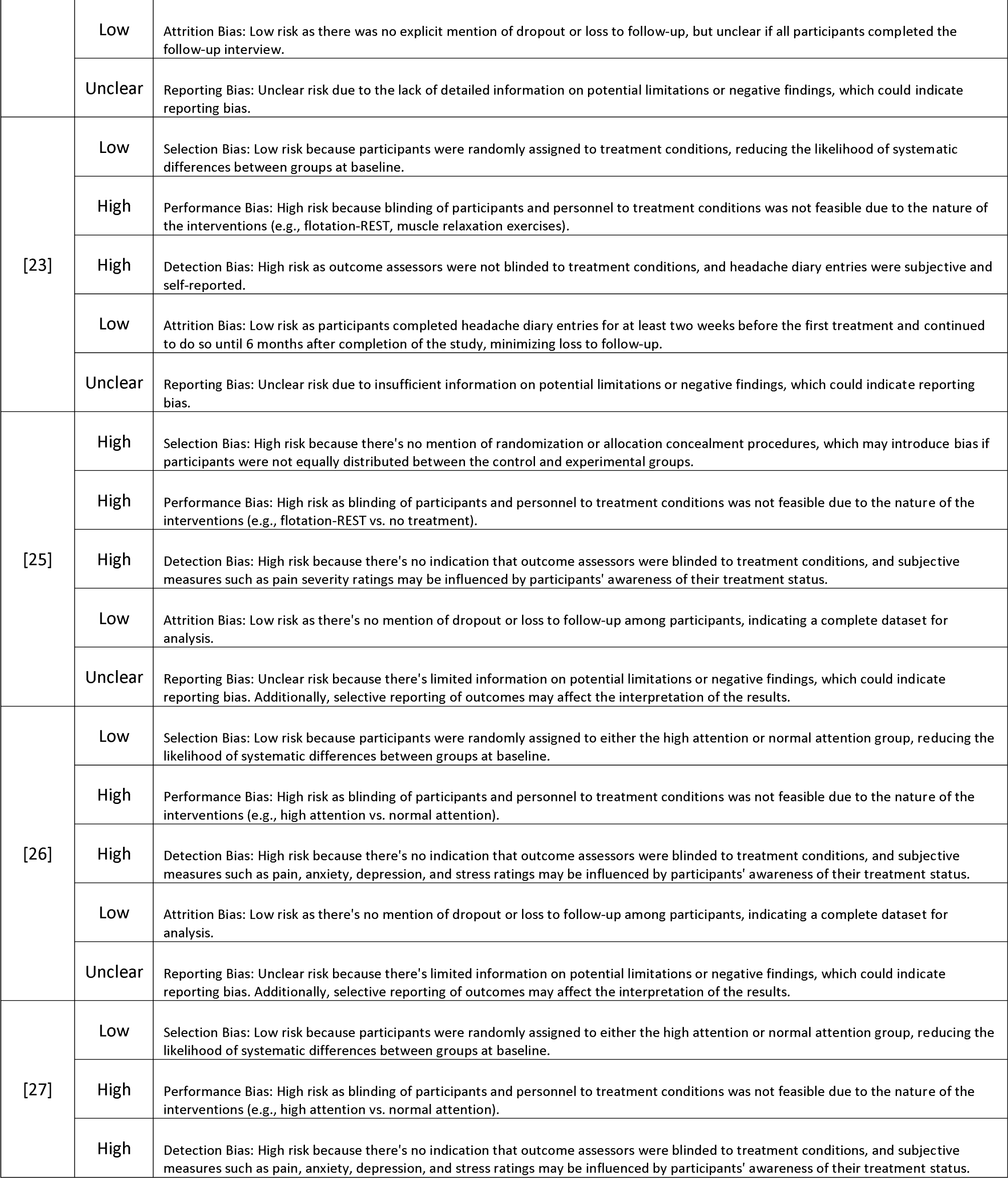
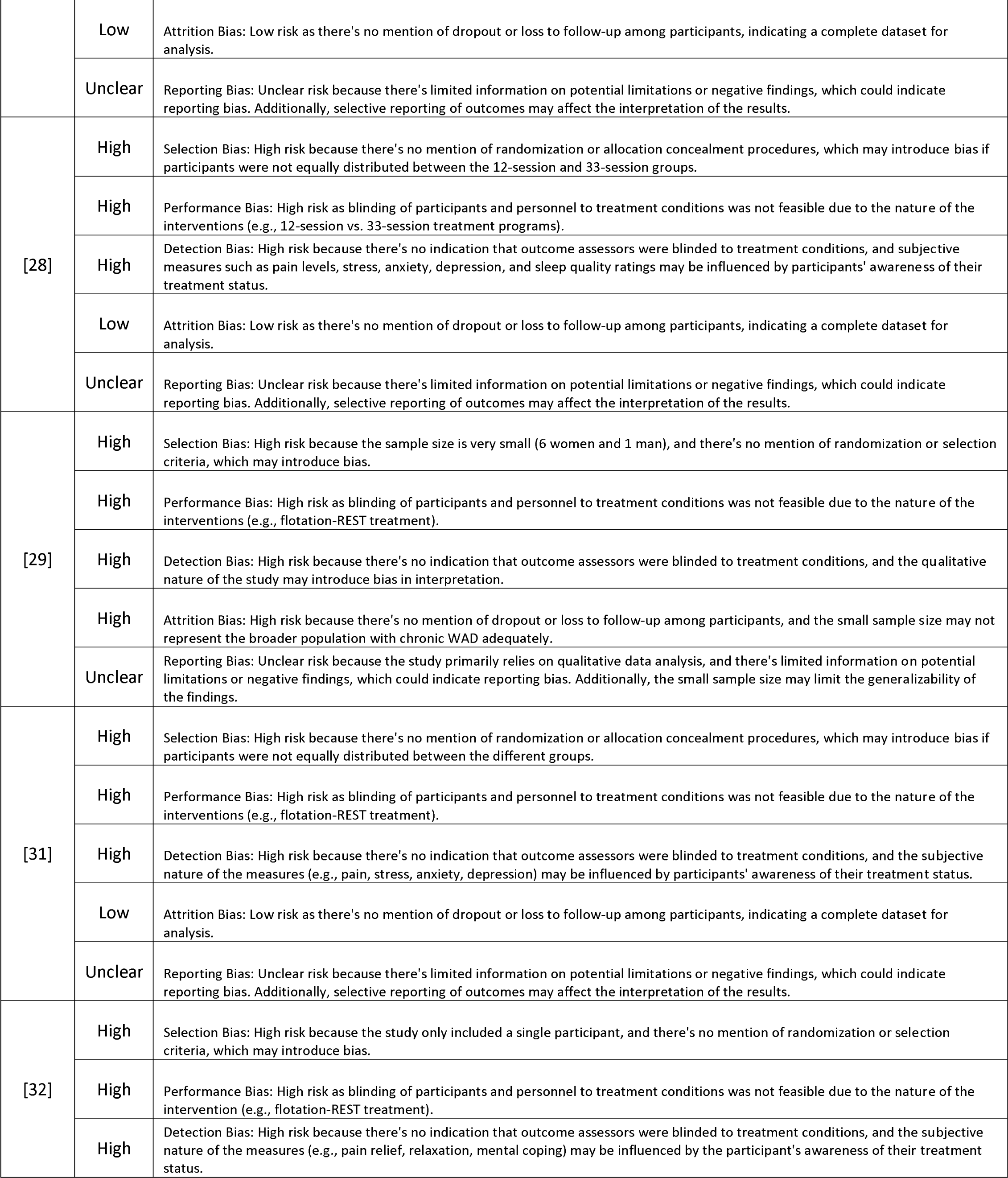
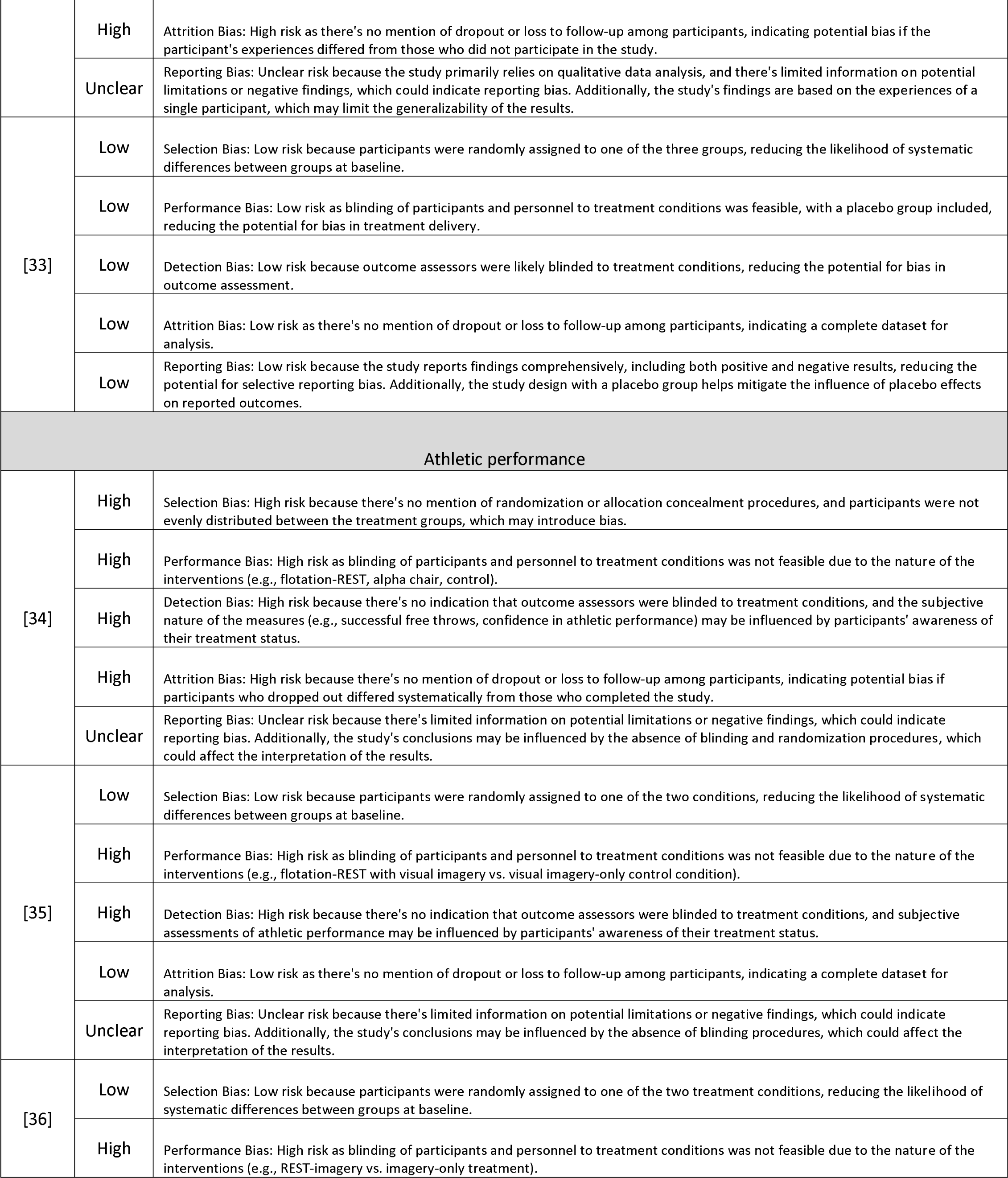
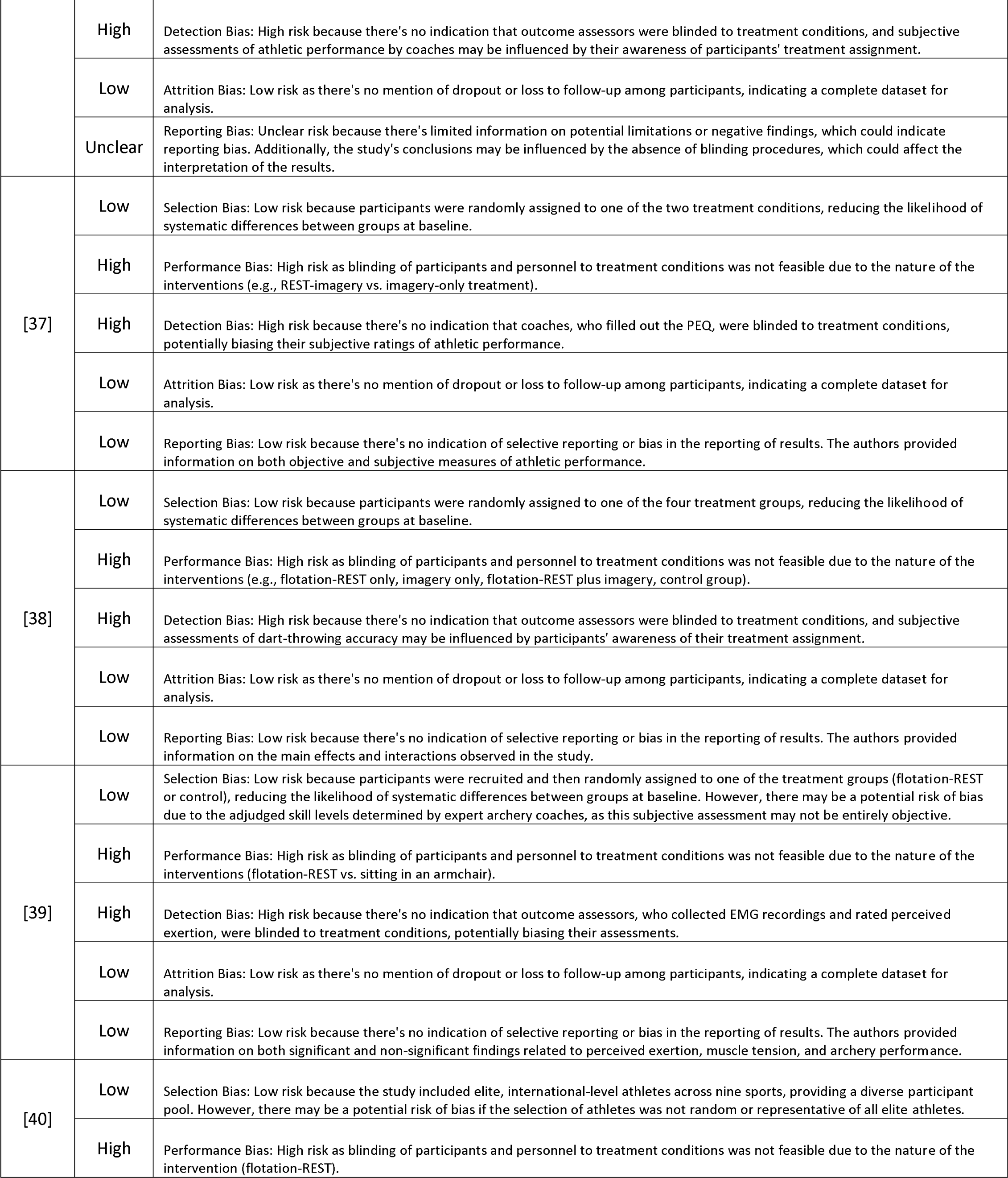
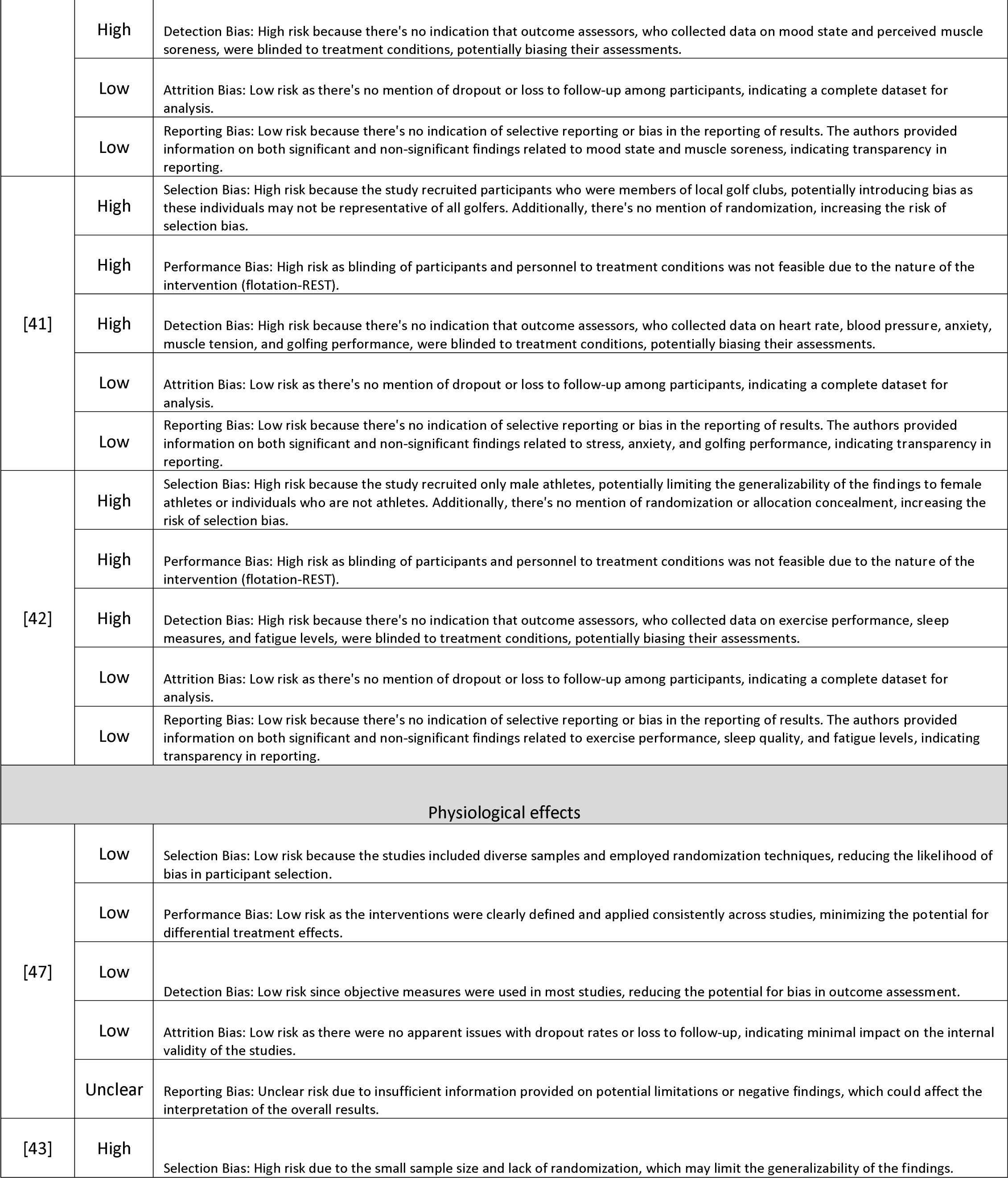
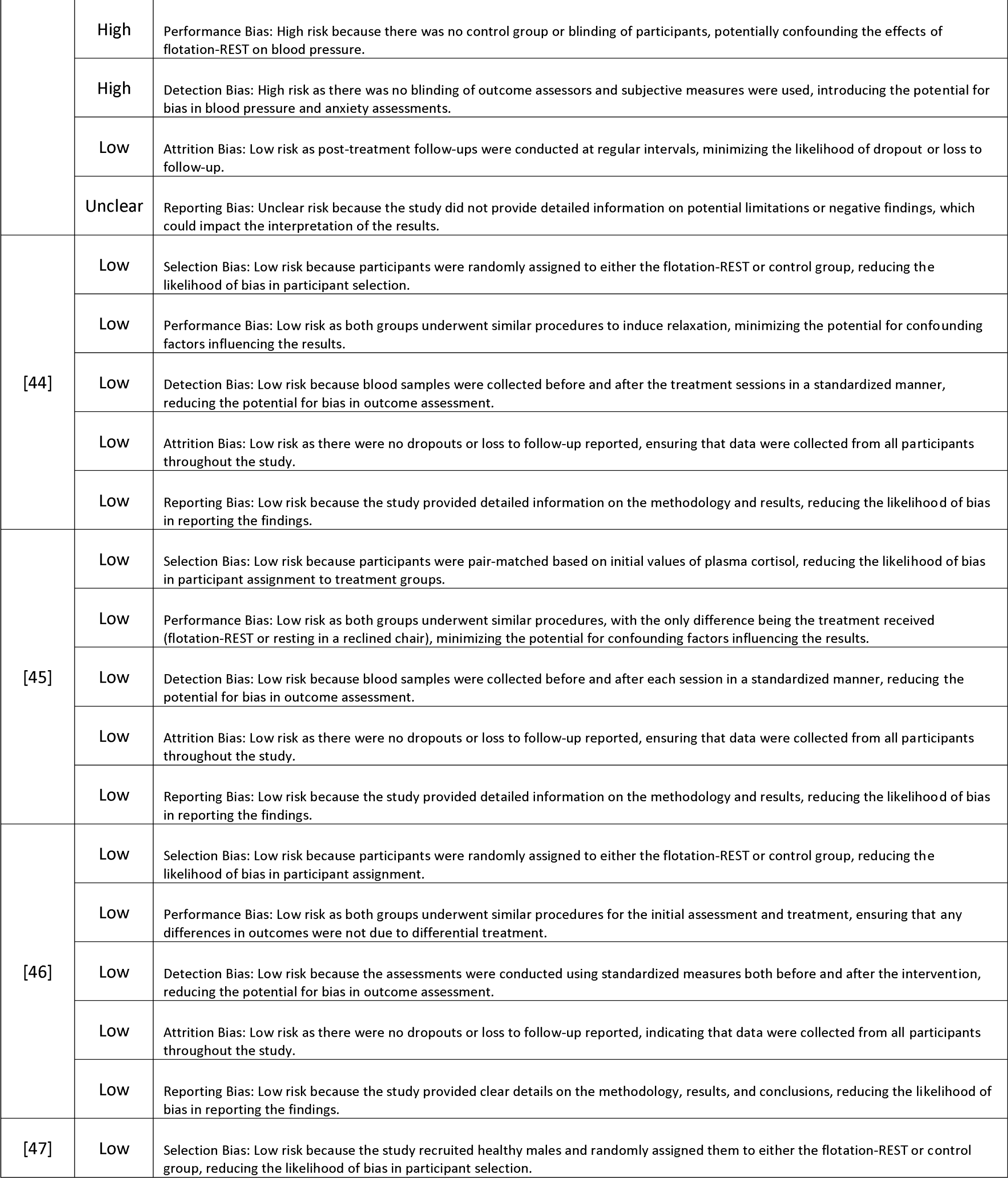
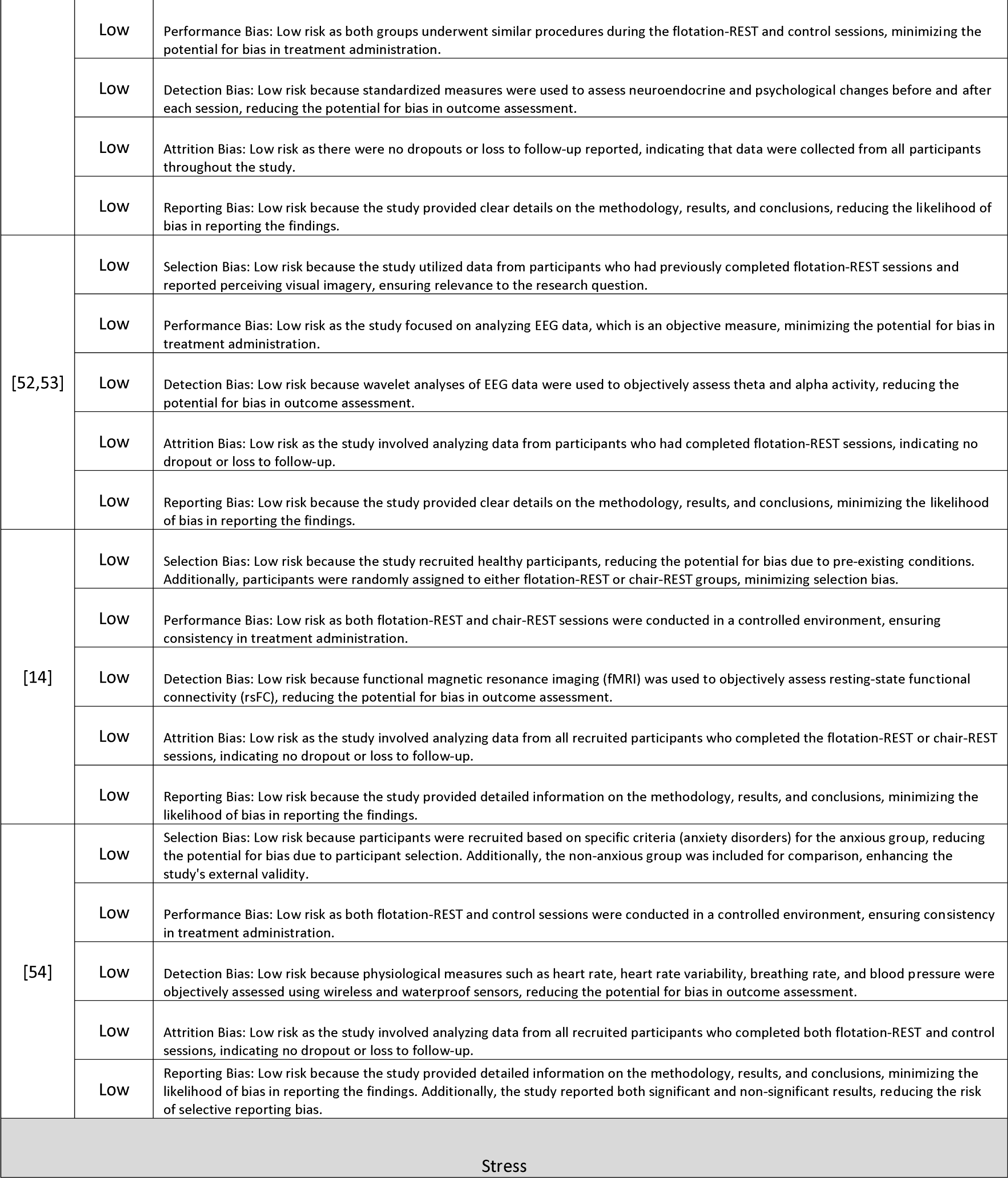
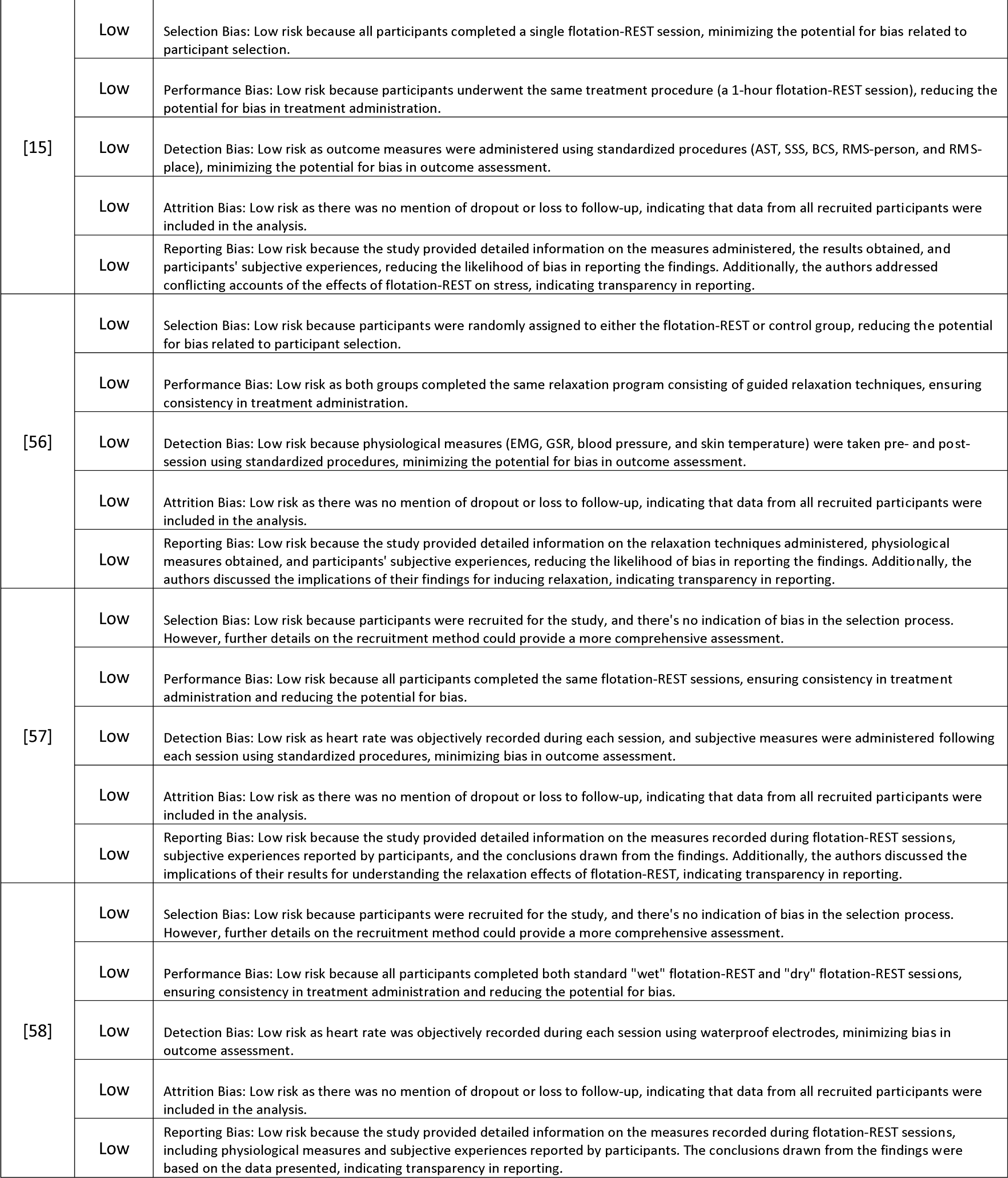
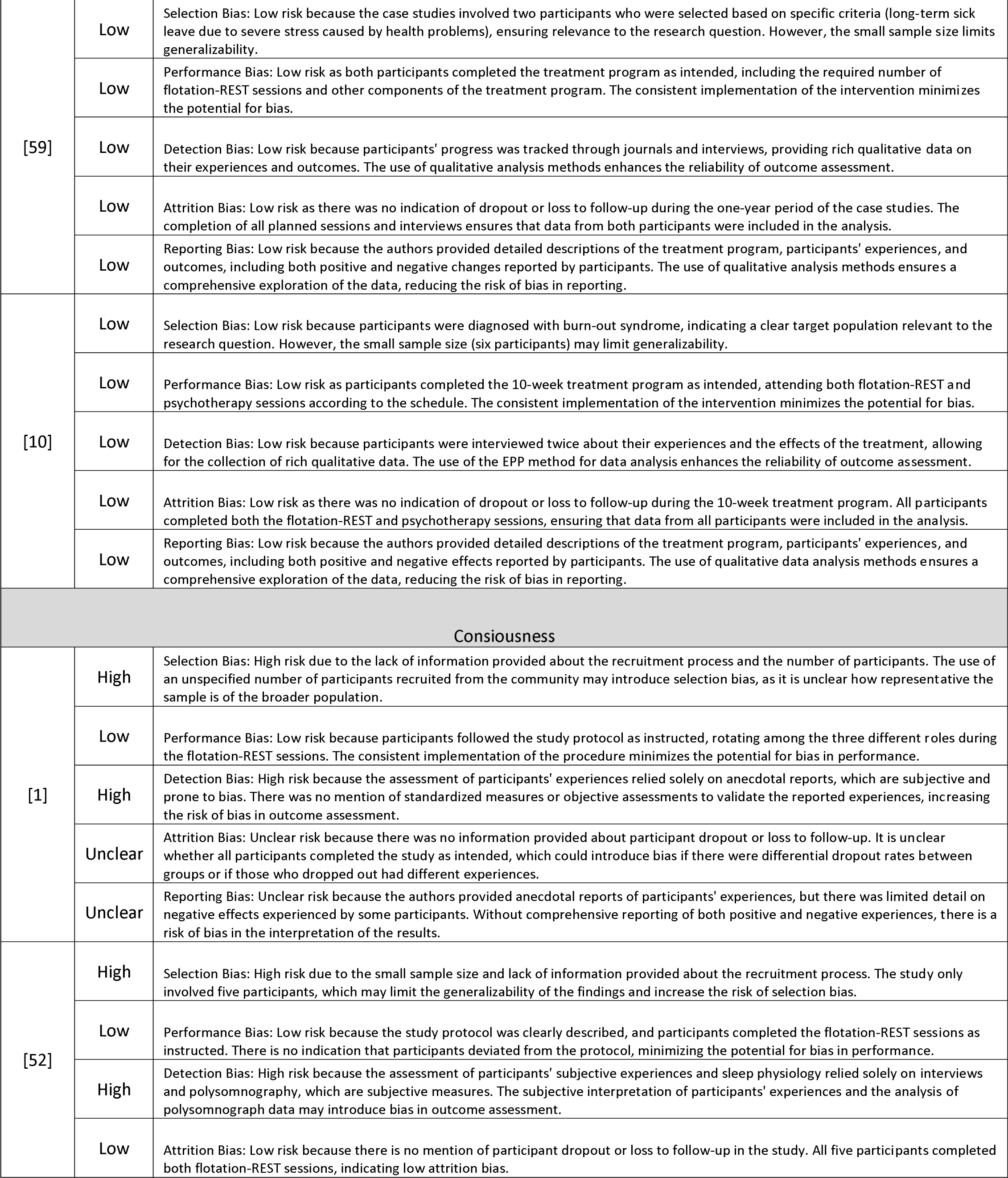
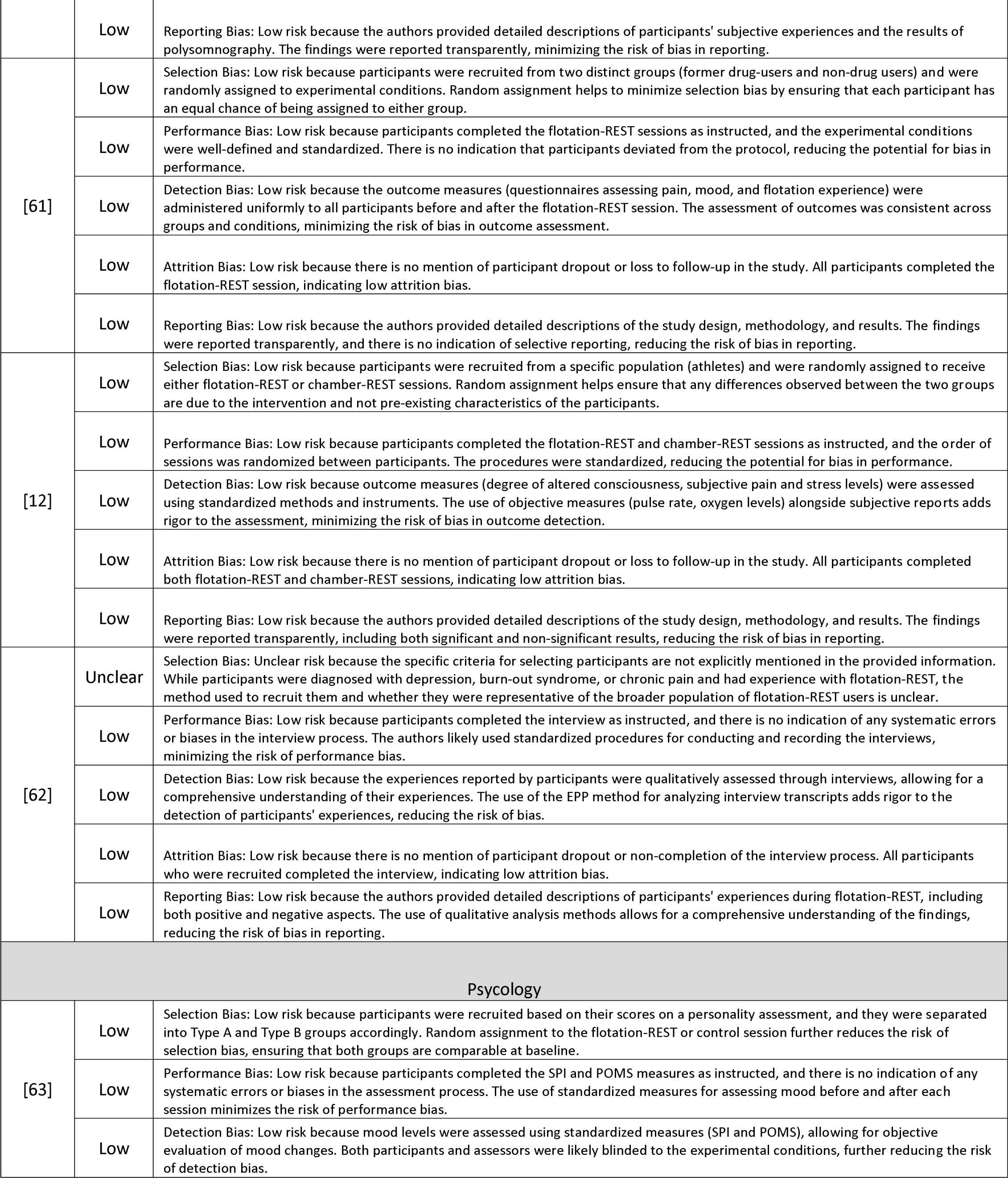
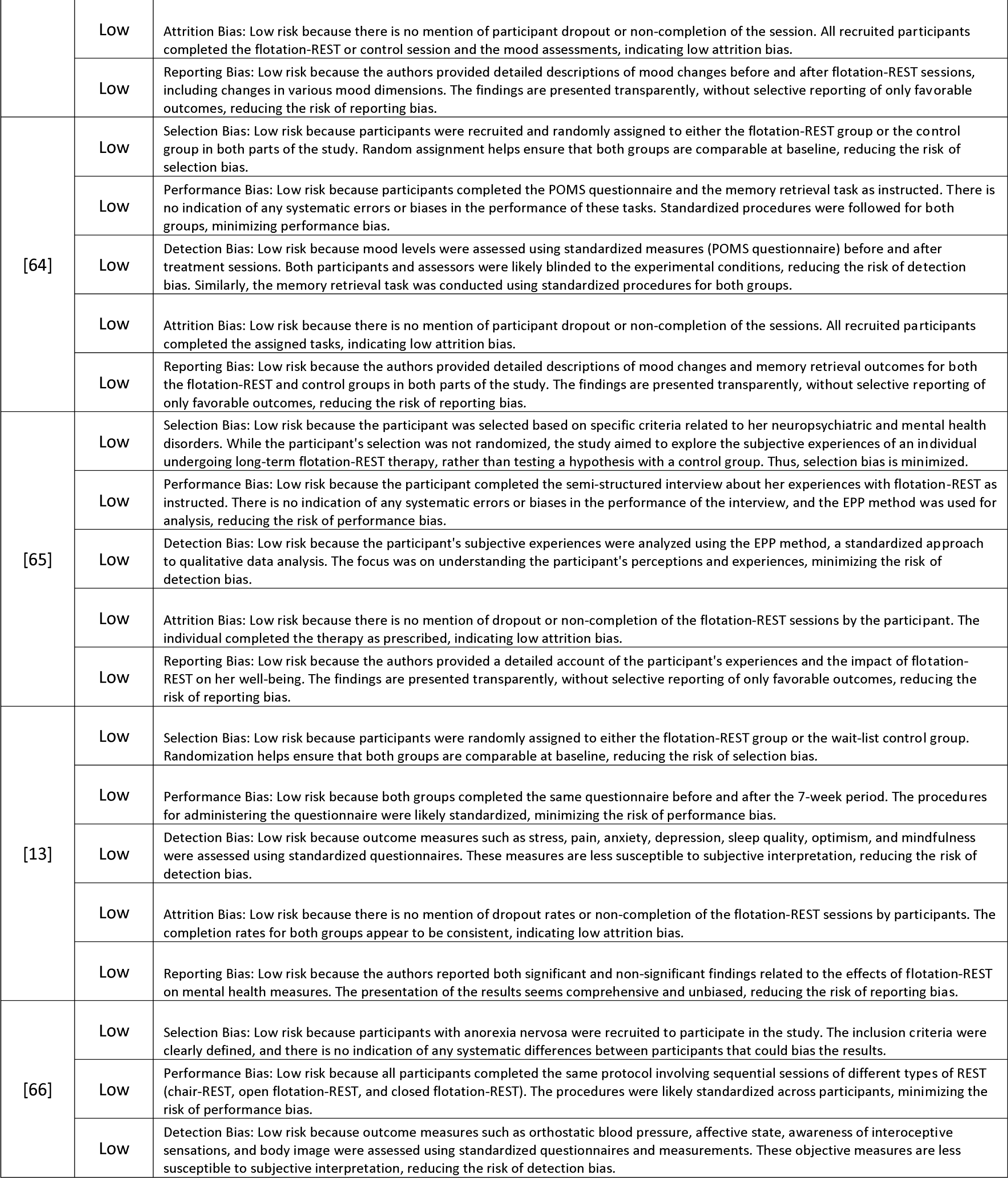
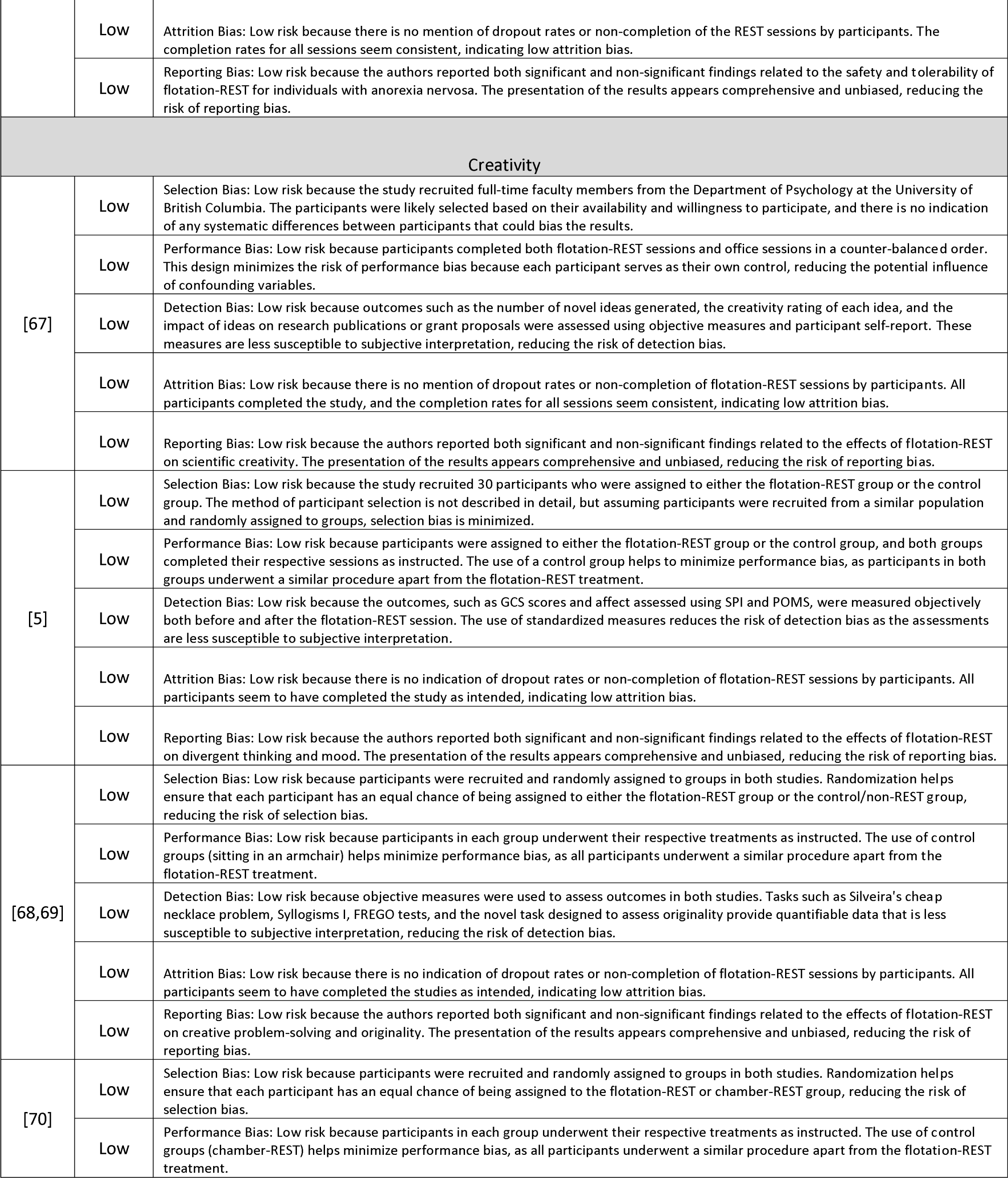
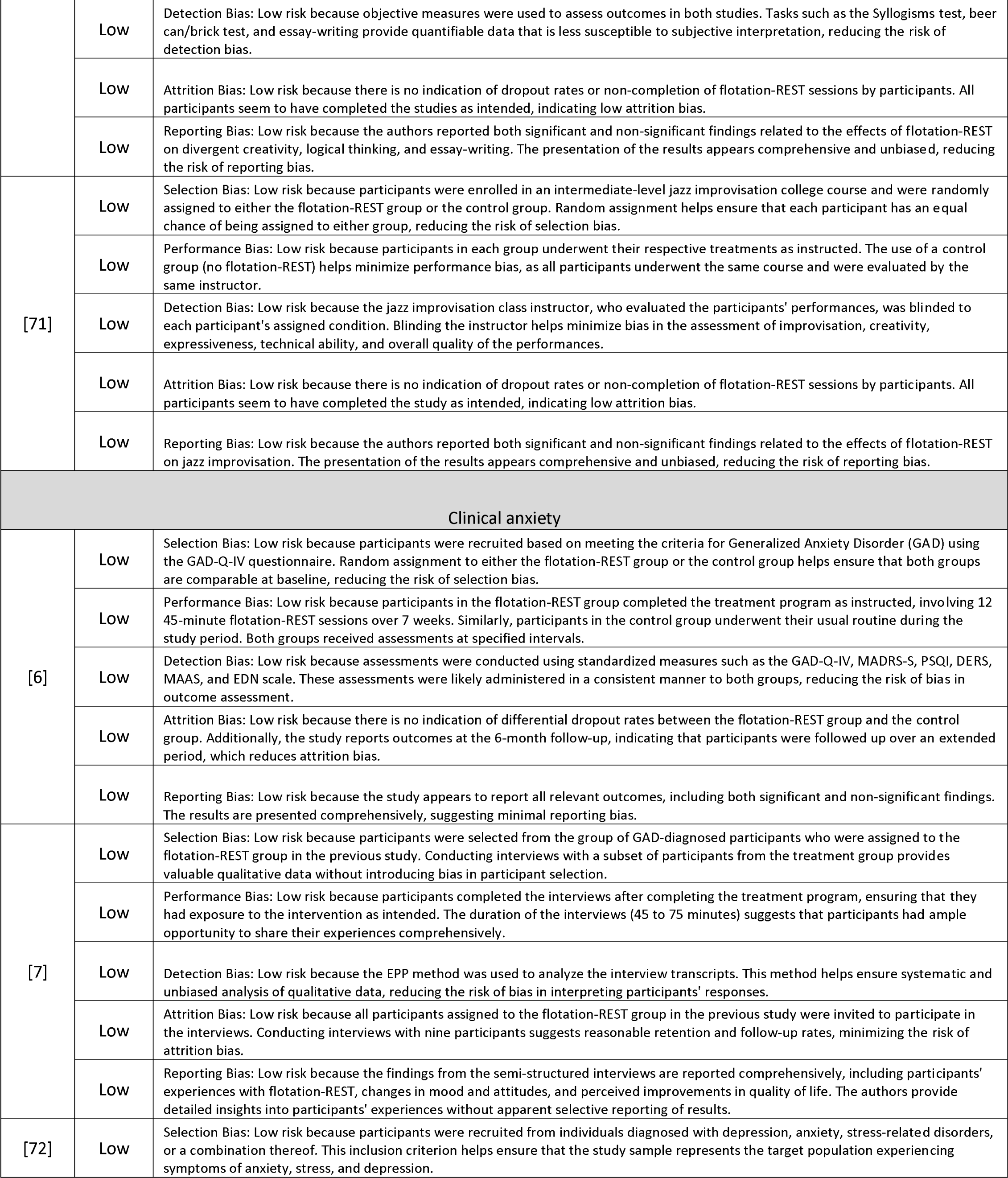
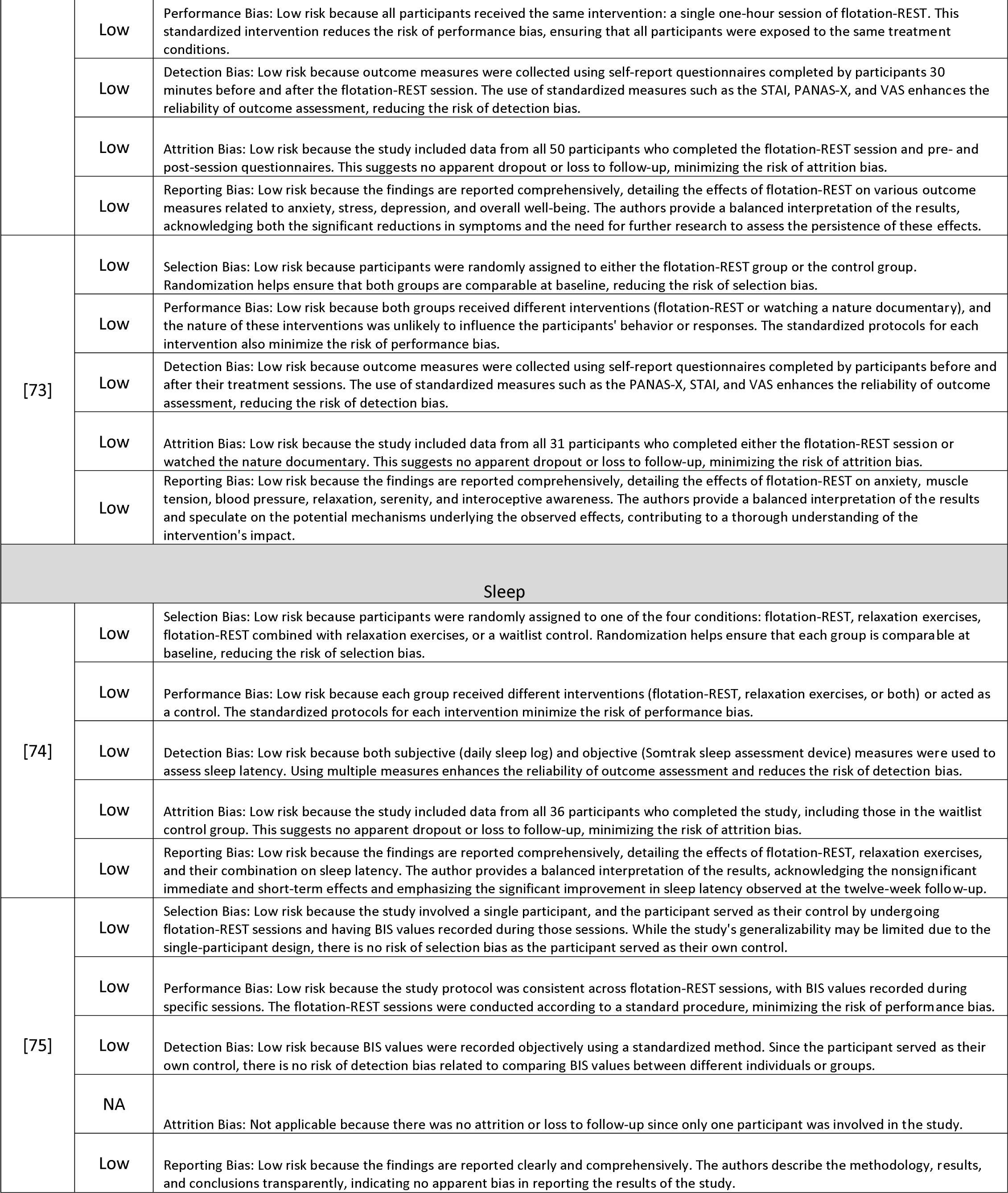
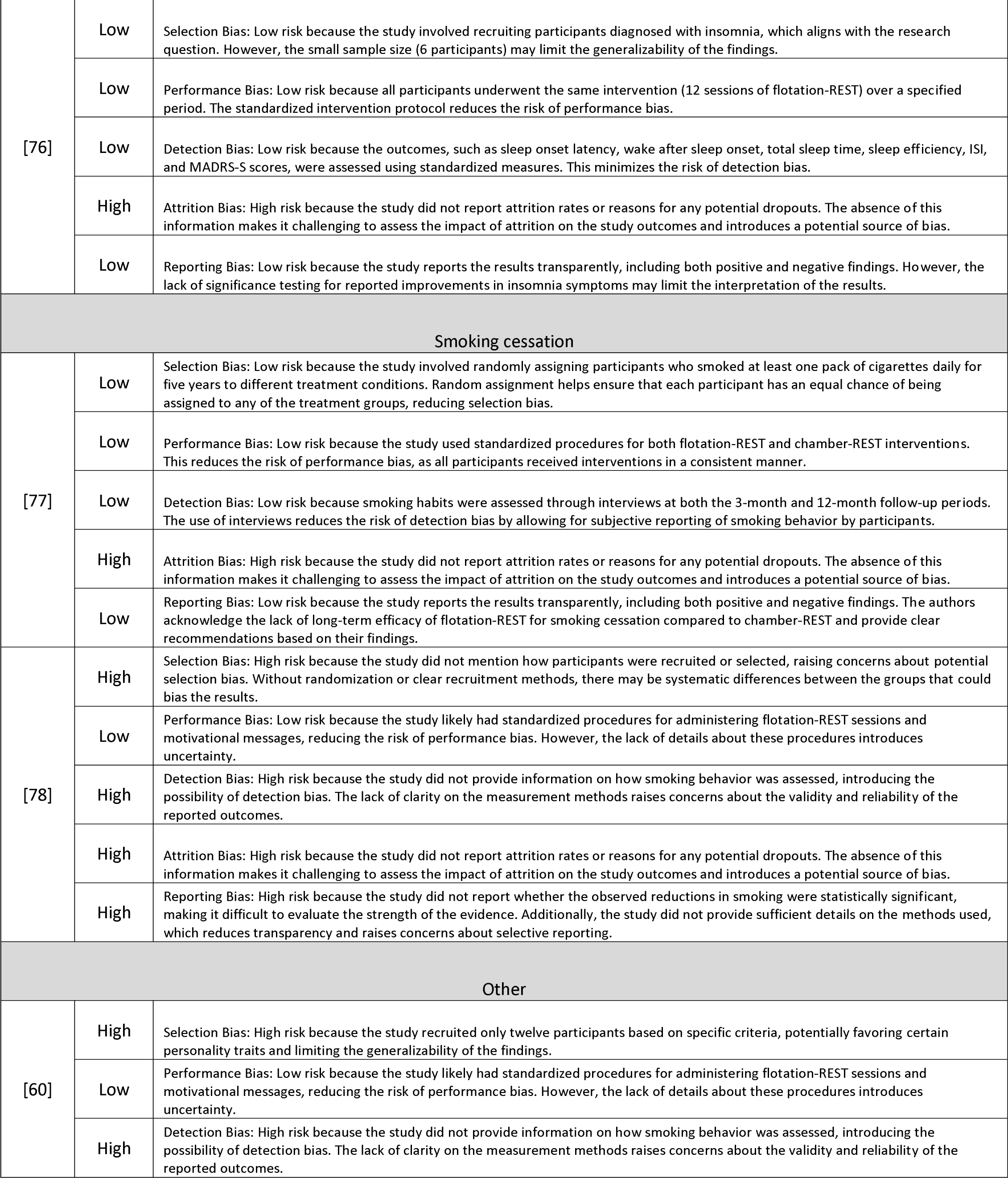
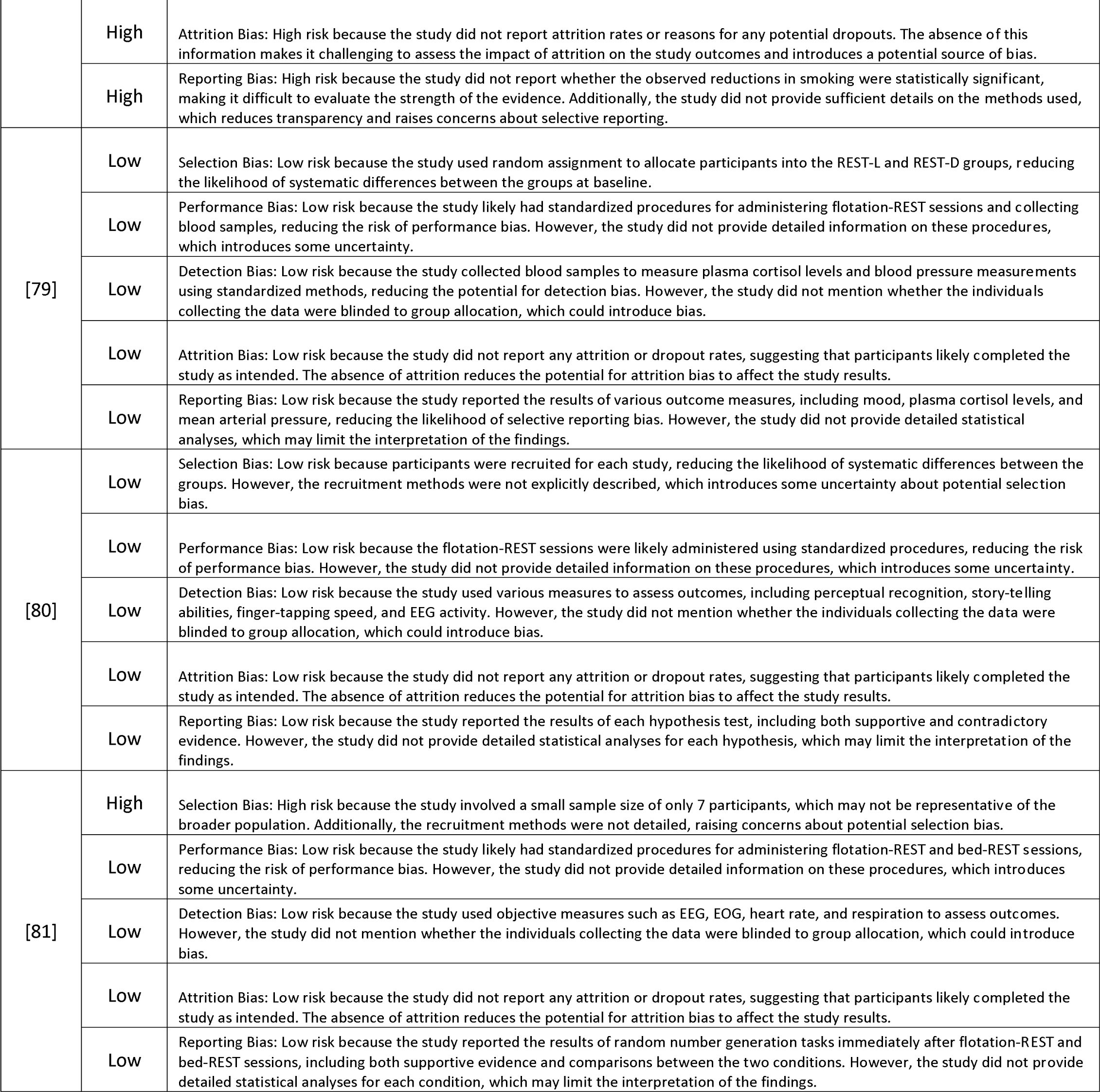
Bias assessment for each study.

**Table 5.** List of 60 reviewed papers in this study.

| Category | Title |
| --- | --- |
| Athletic performance | Effects of flotation restricted environmental stimulation on intercollegiate tennis performance [35] |
| Athletic performance | Enhancing perceptual-motor accuracy through flotation REST [38] |
| Athletic performance | Flotation REST and imagery in the improvement of athletic performance [34] |
| Athletic performance | Flotation REST and imagery in the improvement of collegiate basketball performance [36] |
| Athletic performance | Flotation REST as a stress reduction method: The effects on anxiety, muscle tension, and performance [41] |
| Athletic performance | Flotation restricted environmental stimulation therapy and napping on mood state and muscle soreness in elite athletes: A novel recovery strategy? [40] |
| Athletic performance | Flotation-restricted environmental stimulation therapy improves sleep and performance recovery in athletes [42] |
| Athletic performance | Primary process in competitive archery performance: Effects of flotation REST [39] |
| Clinical anxiety | Characterizing the experiences of flotation-REST (Restricted Environmental Stimulation Technique) treatment for generalized anxiety disorder (GAD): A phenomenological study [7] |
| Clinical anxiety | Examining the short-term anxiolytic effect of flotation-REST [72] |
| Clinical anxiety | Promising effects of treatment with flotation-REST (restricted environmental stimulation technique) as an intervention for generalized anxiety disorder (GAD): A randomized |
| Clinical anxiety | The elicitation of relaxation and interoceptive awareness using flotation therapy in individuals with high anxiety sensitivity [73] |
| Consciousness | Altered consciousness in flotation-REST and chamber-REST: Experience of experimental pain and subjective stress [12] |
| Consciousness | A study on polysomnographic observations and subjective experiences under sensory deprivation [52] |
| Consciousness | Experiments in solitude in maximum achievable physical isolation with water suspension of intact healthy persons [1] |
| Consciousness | Sensory isolation in flotation tanks: Altered states of consciousness and effects on well-being [62] |
| Consciousness | The experience of flotation-REST as a function of setting and previous experience of altered state of consciousness [61] |
| Creativity | Creativity enhancement through flotation isolation [5] |
| Creativity | Effects of flotation REST on creative problem solving and originality [68] |
| Creativity | Effects of flotation- versus chamber-restricted environmental stimulation technique (REST) on creativity and realism under stress and non-stress conditions [70] |
| Creativity | Enhancement of scientific creativity by flotation REST (Restricted Environmental Stimulation Technique) [67] |
| Creativity | The effect of the flotation version of restricted environmental stimulation technique (REST) on jazz improvisation [71] |
| Other | Enhancement of randomness by flotation REST (restricted environmental stimulation technique) [81] |
| Other | Explaining the effects of stimulus restriction: Testing the dynamic hemispheric asymmetry hypothesis [80] |
| Other | Profound experimental sensory isolation [60] |
| Other | The presence or absence of light during flotation [79] |
| Pain | Behavior change and pain relief in chronic whiplash associated disorder Grade IV using flotation restricted environmental stimulation technique: A case report [32] |
| Pain | Chronic whiplash-associated disorders and their treatment using flotation-REST (Restricted Environmental Stimulation Technique) [29] |
| Pain | Effects of flotation REST (Restricted Environmental Stimulation Technique) on stress related muscle pain: Are 33 flotation sessions more effective than 12 sessions? [28] |
| Pain | Effects of flotation-rest on muscle tension pain [11] |
| Pain | Effects of flotation-restricted environmental stimulation technique on stress-related muscle pain: What makes the difference in therapy – attention-placebo or the relaxation response? |
| Pain | Eliciting the relaxation response with the help of flotation-REST (Restricted Environmental Stimulation Technique) in patients with stress-related ailments [27] |
| Pain | Flotation restricted environmental stimulation therapy for chronic pain: A randomized clinical trial [33] |
| Pain | Progressive muscle relaxation and restricted environmental stimulation therapy for chronic tension headache: A pilot study [23] |
| Pain | REST-assisted relaxation and chronic pain [22] |
| Pain | Treating stress-related pain with the flotation restricted environmental stimulation technique: are there differences between women and men? [31] |
| Physiology | A controlled investigation of right hemispheric processing enhancement after restricted environmental stimulation (REST) with flotation [46] |
| Physiology | Effects of relaxation associated with brief restricted environmental stimulation therapy (REST) on plasma cortisol, ACTH, and LH [44] |
| Physiology | Exploring the acute cardiovascular effects of flotation-REST [54] |
| Physiology | Neuroendocrine and psychological effects of restricted environmental stimulation technique in a flotation tank [47] |
| Physiology | Quantitative characteristics of alpha and theta EEG activities during sensory deprivation [53] |
| Physiology | Restricting environmental stimulation influences levels and variability of plasma cortisol [45] |
| Physiology | Taking the body off the mind: Decreased functional connectivity between somatomotor and default-mode networks following flotation-REST [14] |
| Physiology | The effect of brief restricted environmental stimulation therapy in the treatment of essential hypertension [43] |
| Psychology | Autobiographical memory and affect under conditions of reduced environmental stimulation [64] |
| Psychology | Beneficial effects of treatment with sensory isolation in flotation-tank as a preventive health-care intervention – a randomized controlled pilot trial [13] |
| Psychology | Quality of life with flotation therapy for a person diagnosed with attention deficit disorder, atypical autism, PTSD, anxiety and depression [65] |
| Psychology | Reduced environmental stimulation in anorexia nervosa: An early-phase clinical trial [66] |
| Psychology | The use of flotation isolation to modify important Type A components in young adult [63] |
| Sleep | A study of flotation-REST (Restricted Environmental Stimulation Therapy) as an insomnia treatment [76] |
| Sleep | Comparison of Bispectral Index™ values during the flotation restricted environmental stimulation technique and results for stage I sleep: A prospective pilot investigation [75] |
| Sleep | The use of flotation rest in the treatment of persistent psychophysiological insomnia [74] |
| Smoking cessation | Flotation REST as a smoking intervention [78] |
| Smoking cessation | Restricted environmental stimulation therapy of smoking: A parametric study [77] |
| Stress | A direct comparison of the 'wet' and 'dry' flotation environments [58] |
| Stress | Case studies on fibromyalgia and burn-out depression using psychotherapy in combination with flotation-REST: Personality development and increased well-being [59] |
| Stress | Is flotation isolation a relaxing environment? [57] |
| Stress | Preventing sick-leave for sufferers of high stress-load and burnout syndrome: A pilot study combining psychotherapy and the flotation tank [10] |
| Stress | The effects of short term flotation REST on relaxation: A controlled study [56] |
| Stress | Water immersion and flotation: From stress experiment to stress treatment [15] |

## 5. Float pod design

Since its original conception in 1954, the float pod has undergone several revisions to its design (Figure 8). The first float pod, created by physician John C. Lilly, required users to be vertically submerged underwater, necessitating the use of cumbersome breathing masks constantly monitored by a safety operator [82]. Although this first prototype already incorporated certain key features that are still used today – namely, a dark, soundproof enclosure filled with body-temperature water – the generally crude design made float pods largely inaccessible. T address this shortcoming, Lilly collaborated with engineer Glenn Perry to create a new design, one in which users could lie horizontally in a buoyant solution highly saturated with Epsom salt [83]. These updated float pods were released in the 1970s and primarily aimed to increase the device’s convenience. Newer models were built to a muc shallower depth and incorporated filtration systems, upgrades that made water maintenance much more efficient. Additionally, these float pods were smaller, lighter, and less costly to build, constructed of materials like fiberglass and acrylic rather than metals. Ultimately, these changes enabled the float pod to be widely disseminated, including for commercial (i.e., non-academic) use. From the 1990s and onwards, the traditional tank structure has bee replaced by a smaller, sleeker enclosed pod. The modern float pod further maximizes user-friendliness by including additional features like temperature control, built-in audio speakers, and updated sanitation systems.

**Figure 8.**
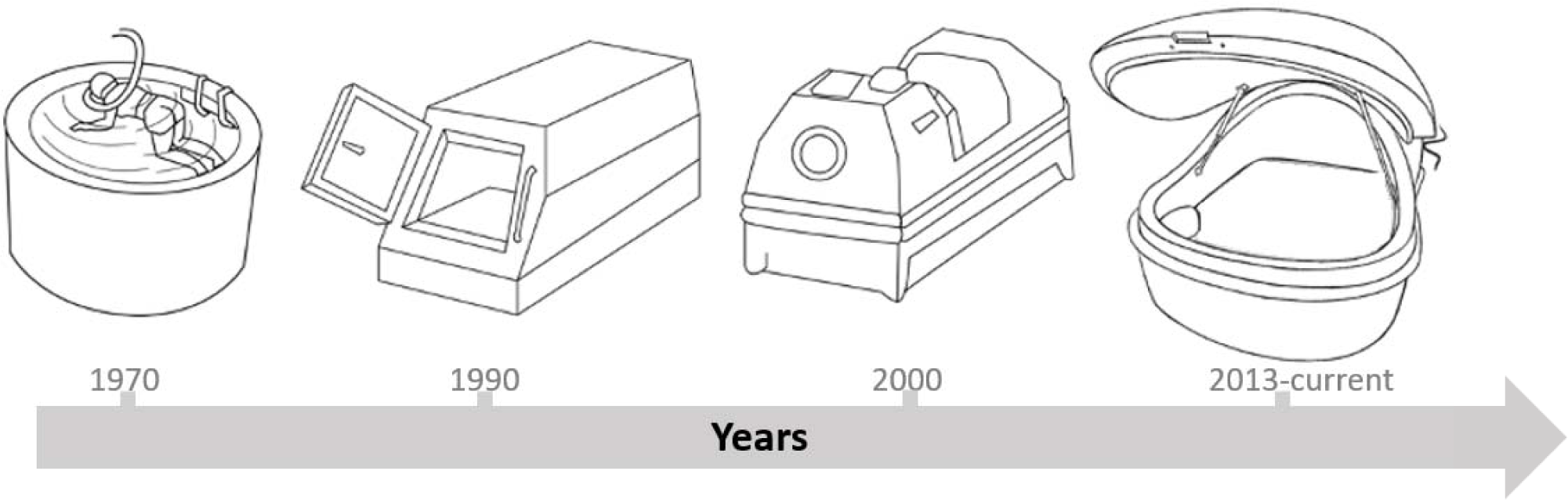
Float pod design over time

Some of the key changes to float pod architecture that have occurred since its conception include:

- Shape: Early float tanks were typically rectangular, while modern float pods are more likely to be rounded or oval in shape. The curved design helps to reduce the amount of external sensory input and create a more cocoon-like environment for users.
- Size: Early float tanks were historically quite large, taking up a significant amount of space in a dedicated flotation room. Modern float pods are typically more compact, allowing them to be installed in a wider range of settings. However, the corrosive nature of the salt used in float pods still necessitates special flooring and easy access to a shower.
- Lighting: Early float tanks typically had no lighting inside the tank, leaving users in complete darkness. Modern float pods may have optional, subtle internal lighting that helps contribute to a calming, relaxing environment without substantially adding to sensory input.
- Acoustics: Early float tanks were often noisy, with audible pumps and filtration systems. Modern float pods are designed to be as quiet as possible, with advanced soundproofing and quiet pumps that create a more peaceful sensory environment, deprived of sound.
- Materials: Early float tanks were typically made of metal, while modern float pods are more likely to be constructed from fiberglass, acrylic, or other high-tech materials. This allows for more precise control over the environment inside the tank and can help to create a more hygienic and low-maintenance experience for users.

## 6. Discussion

Flotation-REST has been shown to have a wide range of potential applications, as demonstrated by the reviewed studies. The 60 studies focused on pain, athletic performance, physiology, stress, consciousness, psychology, creativity, clinical anxiety, sleep, smoking cessation, and other areas. The most common area of application is pain management, with 10 studies (16.7%) focusing on this area. These studies suggest that flotation-REST can be an effective treatment for chronic pain with a variety of origins. Another area of interest is athletic performance, with eight studies (13.3%) investigating the potential benefits of flotation-REST on physical performance and recovery. These studies suggest that flotation-REST may improve certain types of athletic performance as well as enhance post-exercise recovery. Aside from the pain and athletic performance categories, evidence supporting the beneficial effects of flotation-REST was found for the physiology (13.3%), stress (10.0%), consciousness (8.3%), psychology (8.3%), and clinical anxiety (6.7%) categories. Mixed effects were found for the categories of creativity (8.3%), sleep (5.0%), and smoking cessation (3.3%). The number of recruited participants in the 60 reviewed studies ranged from 1 to 99, with an average of 30.6±25.4 participants per study. The methods of treatment delivery for flotation-REST vary, including single or multiple sessions, with session lengths ranging from 35 to 200 minutes. The most common method of treatment delivery reported in the reviewed studies was randomization, followed by pre/post measurements and semi-structured interview designs. The questionnaires used in the reviewed studies to evaluate the effects of flotation-REST were diverse and varied in terms of the psychological and physiological factors they measured. However, a common trend was the use of subjective self-report measures, such as visual analog scales and questionnaires, to assess the effects of flotation-REST. It is important to note that there were limitations in the reviewed studies, including small sample sizes and a lack of control groups, in some cases. Additionally, the studies varied in terms of the type of float pod used, such as traditional float tanks or newer, more advanced models. Overall, however, the reviewed studies suggest that flotation-REST may yield benefits for both physical and mental health. Further research is needed to fully understand the mechanisms of action and potential limitations of this therapy, but the current evidence supports the use of flotation-REST as a complementary or alternative treatment in various areas of health and well-being.

While flotation-REST has shown promise in a range of applications, there are some limitations to consider. First, flotation-REST may not be suitable for everyone. Individuals with certain medical conditions or physical limitations (e.g., open wounds, claustrophobia, severe skin conditions like psoriasis) may not be able to comfortably use flotation tanks, so consulting with a medical professional before participating in flotation-REST is recommended. Second, while flotation-REST can be a relaxing and rejuvenating experience, it is not a substitute for medical or psychological treatment. Flotation-REST should be used as part of a comprehensive approach to health and wellness, rather than as a sole treatment for a specific condition. Finally, cost can be a limitation for some individuals, as flotation-REST can be relatively expensive. Though both commercial float pod facilities and at-home float pods may enjoy an economy of scale if flotation becomes more mainstream.

## 7. Limitations of this review

Some limitations of the current review should be noted. First, only a handful of studies have been published in certain categories of float pod application, so our review should not be used to conclude that flotation-REST is effective for certain understudied applications such as sleep or smoking cessation. The relatively small number of studies available for review limits the generalizability of certain findings and highlights the need for additional research in this area.

Further, the potential for publication bias in the existing literature cannot be ruled out. Researchers often submit predominantly positive results for publication due to a highly competitive publishing landscape. Thus, it is possible that studies reporting negative results were not published and thereby were not included in this review. To increase the likelihood that a negative result will be published, future research should aim to implement study designs with adequate sample sizes, appropriate control groups, and long-term follow-up. Additionally, the development of standardized outcome measures would help to increase comparability across studies and enable more robust meta-analyses.

Despite these limitations, we hope that this review can inform future research on flotation-REST. In particular, this review may serve as a helpful tool for identifying current knowledge gaps in the flotation-REST literature, informing study design, and devising new applications for flotation-REST.

## 8. Future research

There is still much to be explored in the field of flotation-REST. Some potential areas for future research include:

- Long-term effects: Many of the studies reviewed in this analysis had relatively short treatment periods, ranging from a single session to several weeks. Thus, the long-term effects of flotation-REST on different conditions remains understudied.
- Mechanisms of action: While flotation-REST has been shown to have various positive effects, the mechanisms behind these effects are not yet fully understood. Future research could explore the physiological and psychological mechanisms underlying flotation-REST, which could help to optimize treatment approaches and potentially identify new therapeutic targets.
- Optimization of treatment parameters: Further research could explore how the duration, frequency, and other parameters of flotation-REST affect treatment outcomes. The variability of the methods used to collect and report results across studies precludes us from the ability to conduct analyses in order to determine the effects of these parameters.
- Diverse populations: The majority of the studies reviewed in this analysis included predominantly female and/or healthy participants, which limits the generalizability of the findings. Future research could explore the effectiveness of flotation-REST in diverse populations, including individuals with different medical conditions, ages, and genders.

In conclusion, flotation-REST has shown promising potential as a therapeutic intervention for a variety of conditions, and further research could help to optimize its use and identify new applications. It is important, however, to consider the limitations of past research on flotation-REST when planning future studies. Addressing these limitations, such as implementing placebo conditions, including objective measures, and studying more diverse samples, can provide more robust and reliable evidence regarding the effectiveness of flotation-REST. We hope this review will constitute a good entry point for those looking to conduct research on flotation-REST and will assist the field to produce high-quality, reproducible results.

## Registration and Protocol

This study was not registered, and a review protocol was not prepared.

## Data Availability

NA

## Acknowledgment

We affirm that this manuscript is an original work, not previously published, and is not under consideration for publication elsewhere. The publication received support from the Intramural Research Program of the NIH, NINDS, and a joint grant from the John Templeton Foundation and the Fetzer Institute. The views expressed herein are solely those of the author(s) and do not necessarily represent the opinions of NINDS, the John Templeton Foundation, or the Fetzer Institute. The authors do not have any competing interests to disclose.

Our sincere appreciation goes to Dr. Amir Raz for his indispensable support and guidance during the initiation of this project.

